# Increased atherogenicity in mood disorders: a systematic review, meta-analysis and meta-regression

**DOI:** 10.1101/2024.09.19.24313956

**Authors:** Ketsupar Jirakran, Abbas F. Almulla, Thapanee Jaipinta, Asara Vasupanrajit, Priabprat Jansem, Chavit Tunvirachaisakul, Elizabet Dzhambazova, Drozdstoj St. Stoyanov, Michael Maes

## Abstract

**Background:** Major depressive disorder (MDD) and bipolar disorder (BD) often coexist with metabolic syndrome. Both are linked to increased atherogenicity and a higher risk of cardiovascular diseases.

**Objectives:** This meta-analysis seeks to evaluate the relationship between atherogenic indices and mood disorders (MDD/BD), while identifying the most effective atherogenic biomarkers for mood disorders.

**Methods:** This study adhered to the Preferred Reporting Items for Systematic Reviews and Meta-Analyses guidelines. We searched electronic databases, including PubMed, Google Scholar, and Web of Science, for articles published up to August 1, 2024.

**Results:** In this meta-analysis, 85 eligible studies (14 on BD and 71 on MDD) were included, covering 70,856 participants: 18,738 patients and 52,118 healthy controls. Patients with mood disorders showed significant increases (p < 0.001) in the Castelli Risk Index 2 (CRI2), Atherogenic Index of Plasma (AIP), and (triglyceride or TG + low-density lipoprotein + very low-density lipoprotein)/(high-density lipoprotein cholesterol or HDL + Apolipoprotein A or ApoA) ratio, but not CRI1 and ApoB/ApoA ratio. Significant lower HDL and lecithin: cholesterol acyltransferase activity, and higher TG levels were observed in mood disordered patients compared with controls. There were no significant differences between MDD and BD patients. Most included studies lacked the most essential information on the inclusion and exclusion of important confounders.

**Conclusions:** AIP is the most effective atherogenicity index for mood disorders. Regular lipid profiling and metabolic syndrome screening are crucial in mood disorders. Early intervention with lipid-lowering therapies is recommended to prevent the worsening of atherogenicity and disease progression.

## Introduction

Mood disorders including major depressive disorder (MDD) and bipolar disorder (BD) are acknowledged as significant contributors to global disability rates (Walker, McGee et al. 2015). According to the World Health Organization (WHO), the current global prevalence of MDD is estimated to be 4.4%. Mood disorders are known to exhibit a significant comorbidity with metabolic syndrome (MetS) (de Melo, Nunes et al. 2017). Mood disorders and MetS are both characterized by a variety of shared pathways, such as lipid peroxidation, low-grade inflammation, increased insulin resistance, atherogenicity, and diminished antioxidant levels (de Melo, Nunes et al. 2017). Additionally, there are robust bidirectional relationships between mood disorders, atherosclerosis, and cardio-vascular disorders (CVD) (Maes, Kubera et al. 2011, Maes, Ruckoanich et al. 2011). For instance, mood disorders and MDD have been linked to an increased risk of atherosclerosis, heart infarction, and stroke, while stroke and heart infarction are associated with an increased incidence of MDD (Maes, Ruckoanich et al. 2011). The comorbidity between mood disorders and CVD is once again rooted in a variety of shared pathways, such as inflammatory, oxidative, and nitrosative pathways, as well as increased atherogenicity (Maes, Ruckoanich et al. 2011).

Lowered high-density lipoprotein cholesterol (HDL) levels, which indicate increased atherogenicity, underpin mood disorders, MetS, atherosclerosis, and CVD (Maes, Ruckoanich et al. 2011). Serum HDL-cholesterol levels are lower in MDD than in healthy controls (Maes, Smith et al. 1997). Furthermore, there are reports of abnormalities in the levels of triglycerides (TG), total cholesterol, and low-density lipoprotein (LDL) cholesterol (Maes, Zhou et al. 2024). Similarly, atherogenicity indices may be elevated in individuals with mood disorders, such as the Castelli risk index 1 (i.e. total cholesterol/HDL), Castelli risk index 2 (LDL-cholesterol/HDL cholesterol), and the atherogenicity index of plasma (TG/HDL-cholesterol) (Huang and Chen 2004, Huang 2005, Lehto, Hintikka et al. 2008, Moreira, Jansen et al. 2017, Jirakran, Vasupanrajit et al. 2023, Khalfan, Campisi et al. 2023).

The first paper to demonstrate an elevated Castelli risk index 1 in MDD was published in 1994 (Maes, Delanghe et al. 1994). Vargas-Nunes et al. (2015) and Vargas et al. (2014) reported an elevated Castelli risk index 2 and atherogenic Index of Plasma (AIP) in individuals with mood disorders (Vargas, Nunes et al. 2014, Nunes, Piccoli de Melo et al. 2015). However, a limited number of research papers on mood disorders have actually measured total cholesterol and HDL-cholesterol and reported the Castelli risk indices and the AIP index. Additionally, the Maes’ laboratory has published that precisely establishing differences in atherogenicity profiles between MDD and controls is only possible in MDD patients who do not have MetS (Maes, Zhou et al. 2024). The inclusion of subjects with MetS obscures all associations between atherogenicity profiles and MDD, the severity of depression and suicidal behaviors.

In addition, it is worth considering whether there are other atherogenic indices that could be more targeted towards mood disorders than the Castelli risk index 1 and 2, and the AIP. One potential index is based on a combination of TG, LDL and related lipoproteins (e.g. very low-density lipoprotein, VLDL) / HDL and apolipoprotein (Apo)A (Maes, Zhou et al. 2024). Another potentially relevant index in MDD and mood disorders is the ApoB / ApoA ratio (Maes, Zhou et al. 2024).

It is also important to consider the effects of free cholesterol which is atherogenic and even neurotoxic (Almulla, Thipakorn et al. 2023, Maes, Zhou et al. 2024). HDL effectively eliminates free cholesterol from the body through its activity in the reverse cholesterol transport (RCT) (Glomset, Norum et al. 1975, Maes, Delanghe et al. 1994, Maes, Smith et al. 1997, Jirakran, Vasupanrajit et al. 2023) and, consequently, plays a crucial role in safeguarding tissues from the harmful effects of free cholesterol and oxidative stress. The RCT relies heavily on the activity of lecithin: cholesterol acyltransferase (LCAT), an enzyme that plays a crucial role in converting free cholesterol into cholesterylesters (Almulla, Thipakorn et al. 2023). The latter are encapsulated into the HDL particles and subsequently removed from the circulation by the liver (Almulla, Thipakorn et al. 2023). In addition to LCAT, ApoA is associated with the HDL particle and contributes to its antioxidant and anti-inflammatory functions. Consequently, the heightened atherogenicity and reduced RCT can result in lipid peroxidation, giving rise to neoantigens like oxidized LDL. This, in turn, triggers autoimmune reactions against oxidized LDL and sets off inflammatory processes (Almulla, Thipakorn et al. 2023). These processes are fundamental to the development of both atherosclerosis and mood disorders, including MDD (Almulla, Thipakorn et al. 2023). LCAT activity can be estimated from measurements of total cholesterol and cholesteryl-esters by computing the esterified cholesterol ratio (Maes, Delanghe et al. 1994).

Nevertheless, the available data does not provide any conclusive evidence regarding the atherogenicity biomarker that is most strongly associated with MDD/BD, nor did it establish the clinical value of using one of the lipid ratios to assess heightened atherogenicity in MDD/BD. Therefore, this systematic review and meta-analysis will investigate the potential association between atherogenicity indices and MDD/BD, as well as explore how the presence of MetS and other confounding factors may impact the interpretation of these indices in MDD/BD. Thus, this study aims to investigate the most appropriate atherogenicity marker for psychiatrists to utilize in clinical practice. This marker may help to estimate atherogenicity and the subsequent heightened risk of atherosclerosis, CVD, heart infarction, and stroke.

## Materials and methods

To ensure a methodologically robust approach in our research, we adhered to significant frameworks including the Preferred Reporting Items for Systematic Reviews and Meta-Analyses (PRISMA) 2020 (Page, McKenzie et al. 2021), and the guidelines for conducting Meta-Analyses of Observational Studies in Epidemiology (Higgins JPT 2019). We analyzed the differences in atherogenicity indices such as the Castelli Risk Index-1 and 2, the AIP, (TG+LDL+VLDL)/(HDL+ApoA), and ApoB/ApoA between mood disorder patients and controls, and between BD and MDD. Additionally, our analysis also included examining the individual’s lipid biomarkers such as total cholesterol, TG, LDL, VLDL, ApoB, HDL, ApoA, and LCAT activity.

### Search Strategy

Our methodology for compiling comprehensive data on lipid biomarkers included an extensive search of electronic databases, specifically PubMed, Google Scholar, and Web of Science, from January to August 1, 2024. The search was conducted using predefined keywords and MeSH terms, as listed in Table 1 of the supplementary electronic file (ESF). To ensure robustness of the included data, we also carefully examined the reference lists from selected studies and relevant prior meta-analyses, aiming to include all relevant research in our review.

**Table 1.**
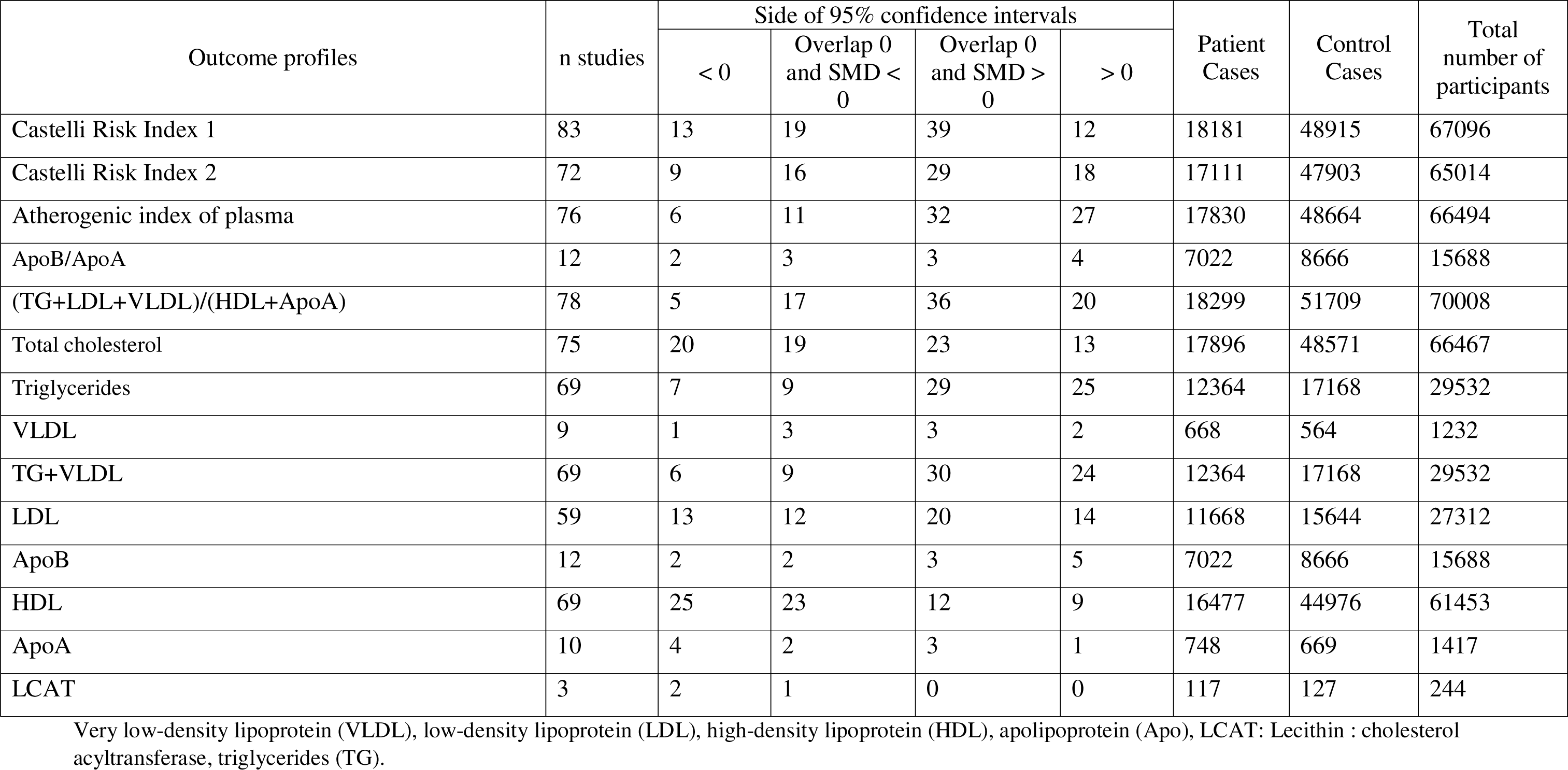
The outcomes and number of patients with major depressive disorder (MDD) and healthy controls and the position of the standardized mean difference (SMD) and 95% confidence intervals with respect to the zero SMD.

### Eligibility criteria

In this meta-analysis, primary inclusion was reserved for studies published in peer-reviewed journals and authored in English, though we also reviewed grey literature and contributions in Thai, French, Spanish, Turkish, German, Italian, and Arabic. We focused on observational, case-control, and cohort studies with controls and MDD patients diagnosed according to DSM or ICD criteria. Studies eligible for inclusion also included those with baseline biomarker assessments in prospective cohort studies or follow-up analyses. Exclusions applied to animal, genetic, and translational research; studies lacking a control group; those employing non-standard mediums such as saliva, hair, whole blood, and platelet-rich plasma; and articles failing to report mean and standard deviation (SD) or standard error (SE) for biomarkers. Nevertheless, for studies missing mean (SD) or (SE) data, we contacted authors requesting this information. In the absence of a response, we calculated mean (SD) from median values using the approach by Wan et al. (Wan, Wang et al. 2014) and an online Mean Variance Estimation tool and estimated mean (SD) from graphical data using Web Plot Digitizer.

### Primary and secondary outcomes

The primary outcome variables of this meta-analysis are the three atherogenicity indices (Castelli risk index 1 and 2, and AIP) in comparison with the (TG+LDL+VLDL)/(HDL+ApoA) ratio and the ApoB/ApoA index in MDD/BD patients versus controls. Additionally, our analysis also included secondary outcomes, namely the individual’s lipid biomarkers such as total cholesterol, TG, LDL, VLDL, ApoB, HDL, ApoA, and LCAT activity.

### Screening and data extraction

The initial screening of studies for potential inclusion in our meta-analysis was conducted by KJ and AA, who reviewed the titles and abstracts according to predetermined inclusion criteria, then proceeded to access the full texts of qualifying papers while excluding those not meeting our criteria. Utilizing a specially designed Excel spreadsheet, they catalogued critical information such as author names, study dates, names of lipid biomarkers, their mean values and SD, the number of participants in patient and control groups, and overall sample sizes. This spreadsheet also included details on study design, type of biological sample (e.g., serum, plasma, CSF, brain tissues, blood cells), psychiatric rating scales, disease stage, average ages (mean and SD), sex distribution, and geographical location of each study. In instances of disagreement, the final adjudication was sought from the senior author (MM).

To assess the methodological quality of included studies, we applied the Immunological Confounder Scale (ICS) (Andres-Rodriguez, Borras et al. 2020), which was adapted for atherogenicity research by the senior author (MM). This tool, detailed in Table 2 of the ESF, comprises the Quality Scale and the Redpoints Scale, both of which have been frequently utilized in prior meta-analyses to evaluate studies on immune and tryptophan catabolite data in individuals with affective disorders (Almulla, Thipakorn et al. 2022, Almulla, Thipakorn et al. 2022, Vasupanrajit, Jirakran et al. 2022). The Quality Scale, which assesses factors such as sample size, control of confounding variables, and sampling duration, scores studies from 0 (lower quality) to 10 (higher quality). The Redpoints Scale aims to identify potential biases in atherogenic biomarkers outcomes and study designs by measuring the degree of control over critical confounding factors, with scores ranging from 0 (highest control) to 26 (no control).

**Table 2.** Results of meta-analyses performed on lipid profiles.

| Outcome feature sets | n | Groups | SMD | 95% CI | z | p | Q | df | p | I<sup>2</sup> (%) | τ<sup>2</sup> | T |
| --- | --- | --- | --- | --- | --- | --- | --- | --- | --- | --- | --- | --- |
| Castelli Risk Index 1 | 83 | Overall | 0.047 | -0.024;0.118 | 1.307 | 0.191 | 670.887 | 82 | <0.0001 | 87.777 | 0.069 | 0.263 |
| Castelli Risk Index 2 | 72 | Overall | 0.161 | 0.083;0.239 | 4.026 | <0.0001 | 674.480 | 71 | <0.0001 | 89.479 | 0.077 | 0.278 |
| Atherogenic index of plasma | 76 | Overall | 0.211 | 0.133;0.289 | 5.321 | <0.0001 | 765.379 | 75 | <0.0001 | 90.201 | 0.082 | 0.287 |
| ApoB/ApoA | 12 | Overall | 0.202 | -0.024;0.427 | 1.749 | 0.080 | 140.907 | 11 | <0.0001 | 92.193 | 0.127 | 0.356 |
| (TG+LDL+VLDL)/HDL+ApoA | 78 | Overall | 0.161 | 0.099;0.223 | 5.102 | <0.0001 | 466.774 | 77 | <0.0001 | 83.504 | 0.044 | 0.210 |
| Total cholesterol | 75 | Overall | -0.099 | -0.201;0.004 | -1.883 | 0.060 | 1458.496 | 74 | <0.0001 | 94.926 | 0.166 | 0.408 |
| Triglycerides (TG) | 69 | Overall | 0.222 | 0.132;0.312 | 4.834 | <0.0001 | 551.120 | 68 | <0.0001 | 87.661 | 0.105 | 0.324 |
| VLDL | 9 | Overall | 0.058 | -0.210;0.326 | 0.422 | 0.673 | 35.731 | 8 | <0.0001 | 77.611 | 0.124 | 0.352 |
| TG+VLDL | 69 | Overall | 0.223 | 0.133;0.313 | 4.851 | <0.0001 | 553.074 | 68 | <0.0001 | 87.705 | 0.105 | 0.324 |
| LDL | 59 | Overall | -0.009 | -0.130;0.112 | -0.144 | 0.885 | 811.862 | 58 | <0.0001 | 92.856 | 0.185 | 0.431 |
| ApoB | 12 | Overall | 0.168 | -0.034;0.370 | 1.631 | 0.103 | 110.271 | 11 | <0.0001 | 90.025 | 0.096 | 0.310 |
| High density lipoprotein * | 69 | Overall | -0.221 | -0.326;-0.116 | -4.135 | <0.0001 | 1216.555 | 68 | <0.0001 | 94.410 | 0.157 | 0.397 |
| ApoA | 10 | Overall | -0.324 | -0.749;0.100 | -1.497 | 0.134 | 128.769 | 9 | <0.0001 | 93.011 | 0.426 | 0.652 |
| LCAT activity | 3 | Overall | -0.863 | -1.689;-0.037 | -2.048 | 0.041 | 16.253 | 2 | <0.0001 | 87.694 | 0.457 | 0.676 |

Very low-density lipoprotein (VLDL), low-density lipoprotein (LDL), high-density lipoprotein (HDL), apolipoprotein (Apo), \*: Indicate a significant difference in group analysis.

### Data analysis

In conducting this meta-analysis, we utilized the CMA V4 software and adhered to the PRISMA guidelines, as outlined in Table 3 of the ESF, requiring a minimum of two studies per lipid biomarker. For calculating the Castelli Risk Index 1 and 2, we assumed dependence in comparison between MDD patients and healthy controls and selected the effect sizes positive for total cholesterol and LDL while negative effect size for HDL. Furthermore, the (TG+LDL+VLDL)/(HDL+ApoA) ratio was calculated by selecting positive effect sizes for TG, LDL, and VLDL and negative for HDL and ApoA. Lastly, the directions of the effect sizes were selected positive for ApoB and negative for ApoA. Group analyses were conducted to compare outcomes between MDD and BD, as well as between studies that included participants with MetS and those that excluded them. If no significant differences were found, the groups were combined. We adopted a random-effects model using constrained maximum likelihood for pooling effect sizes, defining statistical significance as a p-value less than 0.05, and expressing effect sizes as standardized mean differences (SMD) with 95% confidence intervals (CIs). SMDs of 0.80, 0.5, and 0.20 were indicative of large, moderate, or small effect sizes, respectively, following Cohen’s classification (Cohen 2013). Heterogeneity was assessed using tau-squared (and secondary by Q, and I^2^ values), while meta-regression was applied to explore the sources of heterogeneity.

**Table 3.** Results on publication bias.

| Outcome feature sets | Fail safe n | Z Kendall's $\tau$ | p | Egger's t test (df) | p | Missing studies (side) | After Adjusting |
| --- | --- | --- | --- | --- | --- | --- | --- |
| Castelli Risk Index 1 | 1.065 | 0.353 | 0.361 | 1.692(81) | 0.047 | 17 (Left) | -0.088 (-0.161;-0.015) |
| Castelli Risk Index 2 | 8.379 | 0.665 | 0.252 | 1.263(70) | 0.105 | 13 (Right) | 0.318 (0.231; 0.405) |
| Atherogenic index of plasma | 12.137 | 0.246 | 0.402 | 0.908(74) | 0.183 | 16 (Left) | 0.084 (0.007;0.162) |
| ApoB/ApoA | 1.084 | 0.754 | 0.225 | 2.445(10) | 0.017 | 3 (Right) | 0.386 (0.008;0.764) |
| (TG+LDL+VLDL)/(HDL+ApoA) | 9.292 | 0.533 | 0.296 | 1.840(77) | 0.034 | 12 (Left) | 0.056 (-0.006;0.120) |
| Total cholesterol | -8.866 | 2.515 | 0.005 | 2.121(73) | 0.018 | 14 (Left) | -0.252 (-0.352;-0.152) |
| Triglycerides (TG) | 11.323 | 0.305 | 0.379 | 0.371(67) | 0.355 | 20 (Left) | 0.010 (-0.085;0.106) |
| VLDL | 1.305 | 1.146 | 0.125 | 1.269(7) | 0.122 | 0 | - |
| TG+VLDL | 11.380 | 0.295 | 0.383 | 0.384(67) | 0.350 | 21 (Left) | 0.001 (-0.094;0.098) |
| LDL | -0.485 | 1.582 | 0.056 | 1.466(57) | 0.073 | 14 (Left) | -0.261 (-0.393;-0.130) |
| ApoB | 0.495 | 0.754 | 0.225 | 2.647(10) | 0.012 | 1 (Left) | 0.086(-0.117;0.289) |
| HDL | -12.485 | 1.786 | 0.036 | 0.080(67) | 0.467 | 15 (Left) | -0.418(-0.527;-0.308) |
| ApoA | -5.281 | 0.357 | 0.360 | 0.289(8) | 0.389 | 3 (Left) | -0.578 (-1.010;-0.146) |
| LCAT activity | -5.133 | 1.044 | 0.148 | 2.320 (1) | 0.129 | 0 | - |

Very low-density lipoprotein (VLDL), low-density lipoprotein (LDL), high-density lipoprotein (HDL), apolipoprotein (Apo).

To verify that the effect estimates are robust, leave-one-out sensitivity analyses were performed. The fail-safe N approach, continuity-corrected Kendall tau, and Egger’s regression intercept were used to assess publication bias; the last two methods yielded one-tailed p-values. The trim-and-fill method was used to account for any missing studies and compute modified effect sizes in cases of asymmetry identified by Egger’s test. Additionally, both observed and imputed missing studies were presented using funnel plots, which were used to visualize tiny study effects.

## Results

### Search results

Using predetermined keywords and MeSH terms, as listed in Table 1 of the ESF, we conducted a systematic search and found 26,593 studies. The PRISMA flow chart presented in Figure 1 gives an overview of the procedure, showing the total number of publications included and excluded. After the first round of screening and eliminating research deemed irrelevant, 25,010 articles were discarded, resulting in a selection of 1,583 studies. 90 research (were judged suitable for inclusion in the meta-analysis after an additional 1,493 publications were eliminated based on our inclusion-exclusion criteria. Furthermore, we omitted 5 studies due to the lack of required data from the analysis. Overall, 85 studies (10 BD, 71 MDD, and 4 including both MDD and BD cohorts) were used to provide data from 70,856 participants, ages ranging from 15 to 78 years, were included in this meta-analysis (Oxenkrug, Branconnier et al. 1983, Bajwa, Asnis et al. 1992, Maes, Delanghe et al. 1994, Olusi and Fido 1996, Maes, Smith et al. 1997, Khalid, Lal et al. 1998, Partonen, Haukka et al. 1999, Sanyal, Chakrabarti et al. 2000, Bilici, Efe et al. 2001, Sevincok, Buyukozturk et al. 2001, Äijänseppä, Kivinen et al. 2002, Ranjekar, Hinge et al. 2003, Ergün, Uguz et al. 2004, Huang and Chen 2004, Karlović, Buljan et al. 2004, Kim and Myint 2004, Nakao and Yano 2004, Huang 2005, Jow, Yang et al. 2006, Sarandol, Sarandol et al. 2006, Onuegbu, Agbedana et al. 2007, Lehto, Hintikka et al. 2008, Cizza, Eskandari et al. 2009, Hamidifard, Fakhari et al. 2009, Kodydková, Vávrová et al. 2009, Sagud, Mihaljevic-Peles et al. 2009, Ahola, Thorn et al. 2010, Das, Malhotra et al. 2010, Heckbert, Rutter et al. 2010, Lehto, Niskanen et al. 2010, Roohafza, Sadeghi et al. 2010, van Reedt Dortland, Giltay et al. 2010, ALİYAZICIOĞLU, DEĞER et al. 2011, Baghai, Varallo-Bedarida et al. 2011, Kotan, Sarandol et al. 2011, Olié, Picot et al. 2011, Ruljancic, Mihanovic et al. 2011, Sadeghi, Roohafza et al. 2011, Sonal Sukreet and Chaturvedi 2011, Hocaoglu, Kural et al. 2012, ERMAN, KARA et al. 2013, Rybka, Kędziora-Kornatowska et al. 2013, Bortolasci, Vargas et al. 2014, Kale, Kale et al. 2014, Palta, Golden et al. 2014, Patra, Khandelwal et al. 2014, Rahiminejad, Moaddab et al. 2014, Scharnholz, Gilles et al. 2014, Vargas, Nunes et al. 2014, Kahl, Schweiger et al. 2015, Liu, Zheng et al. 2015, Nunes, Piccoli de Melo et al. 2015, Ormonde do Carmo, Mendes-Ribeiro et al. 2015, Peng, Xiang et al. 2016, Tunçel Ö, Akbaş et al. 2016, Akgün, Köken et al. 2017, Ekinci and Ekinci 2017, Messaoud, Mensi et al. 2017, Moreira, Jansen et al. 2017, Peng, Zhong et al. 2017, Baghai, Varallo-Bedarida et al. 2018, Enko, Brandmayr et al. 2018, Segoviano-Mendoza, Cárdenas-de la Cruz et al. 2018, Tunçel Ö, Sarısoy et al. 2018, Al-Amarei, Rasheed et al. 2019, Eidan, Al-Harmoosh et al. 2019, Hui, Yin et al. 2019, Péterfalvi, Németh et al. 2019, Su, Li et al. 2019, Wagner, Musenbichler et al. 2019, Karadeniz, Yaman et al. 2020, Zhang, Yang et al. 2020, Guidara, Messedi et al. 2021, Kennedy, Islam et al. 2021, Vaghef-Mehrabani, Izadi et al. 2021, Draghici 2022, Kasak, Ceylan et al. 2022, Shapiro, Kennedy et al. 2022, Wei, Wang et al. 2022, Jirakran, Vasupanrajit et al. 2023, Khalfan, Campisi et al. 2023, Chen, Sun et al. 2024, Maes, Zhou et al. 2024, Qi, Wang et al. 2024).

**Figure 1.**
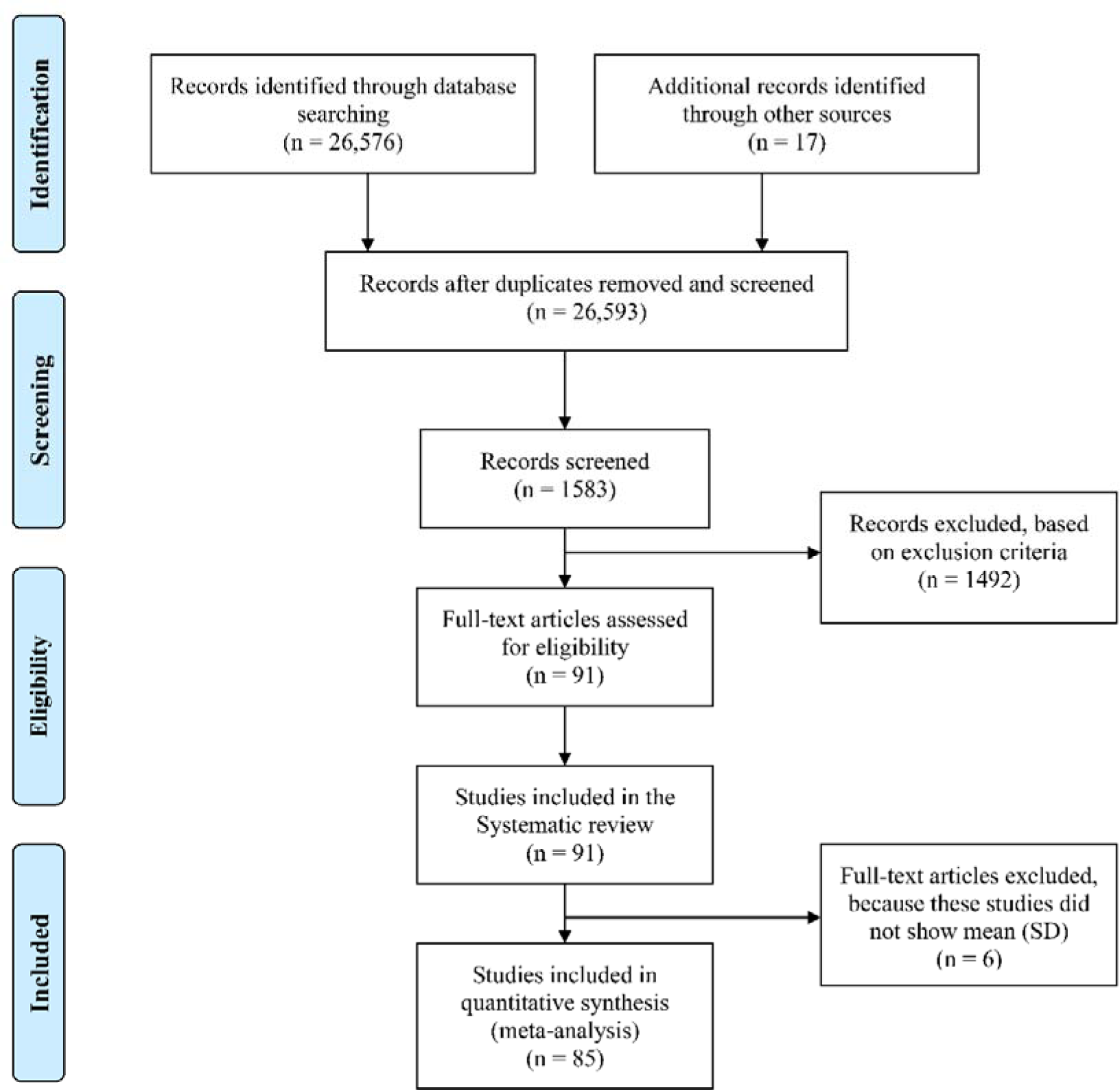
The PRISMA flow chart.

These participants included 52,118 healthy controls and 18,738 patients (6,622 BD and 12,425 MDD). Turkey, India, and China led the way in terms of the geographical contribution of studies, with 14, 7, and 9 articles, respectively. Additionally, as indicated in ESF, Table 5, the USA, Brazil, Finland, Germany, and Iran each published five research papers, and a wide range of other nations contributed one to four studies each. ESF, Table 5 provides specifics on the quality and redpoint scores determined by the study and expressed as median (min–max).

### Primary outcome variables

#### Atherogenic indices

No significant differences were identified between MDD and BD patients in any of the group analyses; hence, both groups were combined. **Table 1** depicts the effect size for the Castelli Risk Index 1 based on 83 studies. **Table 2** and ESF, Figure 1 reveal no significant difference in the Castelli Risk Index 1 between MDD/BD patients and healthy controls. Nonetheless, the publication bias analysis (**Table 3**) uncovered 17 missing studies on the funnel plot’s left side, and incorporating these missing studies shifted the SMD value (-0.088) to a significant decrease.

The effect size for Castelli Risk Index 2 was determined using data from 72 studies. Among these, 9 studies demonstrated CIs entirely below zero, and 18 studies exhibited CIs exclusively above zero. Most of the studies (n=45) had overlapping CIs with varied outcomes; 16 indicated negative SMD values, while 29 documented positive SMD values, as shown in Table 1. Table 2 and **Figure 2** indicate that Castelli Risk Index 2 was significantly higher in patients with MDD/BD than healthy controls, albeit the SMD value was modest. There was no publication bias (see Table 3).

**Figure 2.**
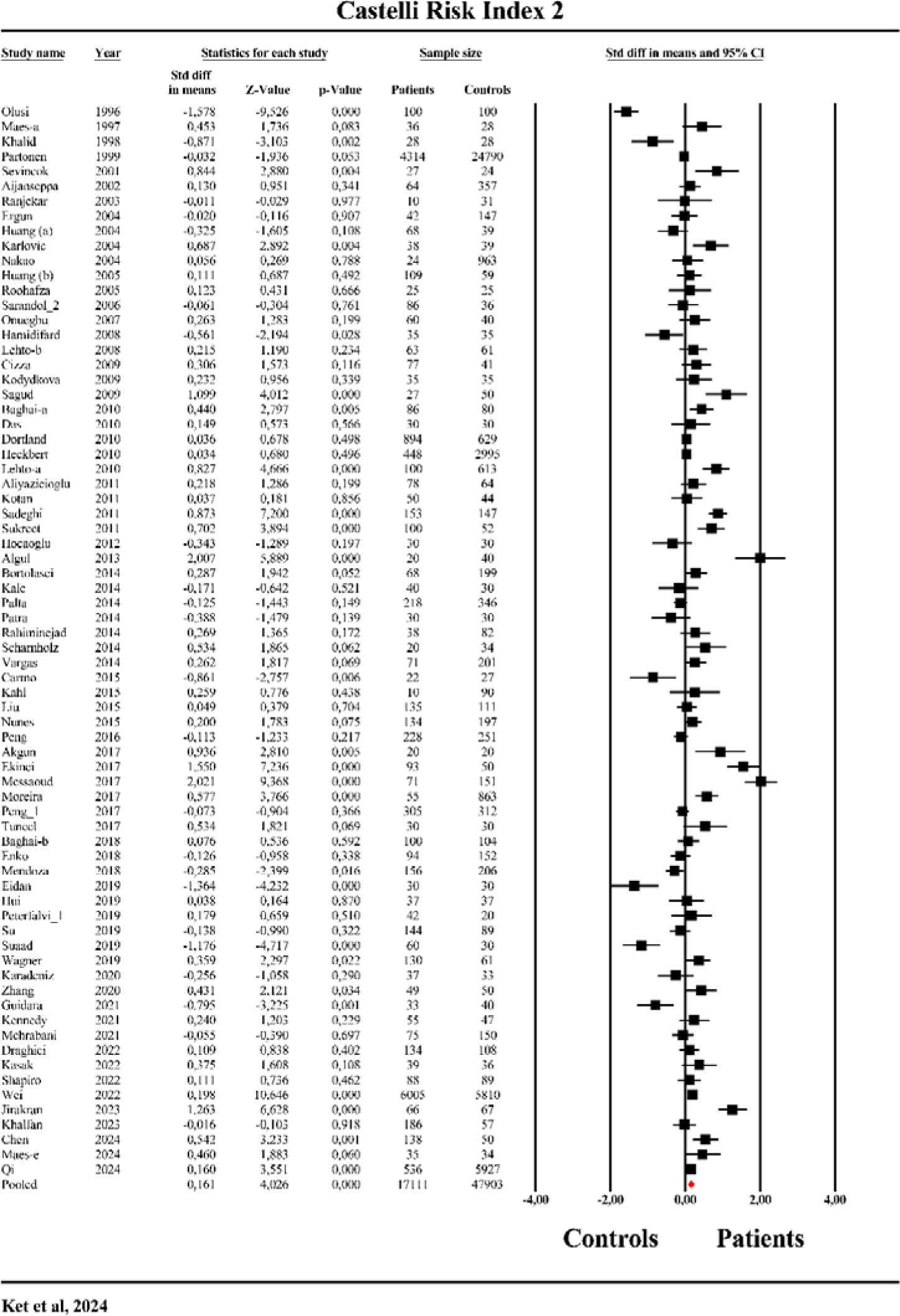
The forest plot of Castelli Risk Index-2 in patients with major depression as compared to healthy controls.

We examined data from 76 studies to assess the effect size of the AIP (TG/HDL ratio). This analysis revealed that 6 studies had CIs entirely below zero, while 27 had CIs exclusively above zero. A significant portion of the research, involving 43 studies, showed overlapping CIs with diverse outcomes: 11 yielded negative SMD values, and 32 reported positive SMD values, as elaborated in Table 1. Table 2 and **Figure 3** illustrate that the TG/HDL ratio was significantly higher in patients with MDD/BD versus healthy controls. There was no publication bias.

**Figure 3.**
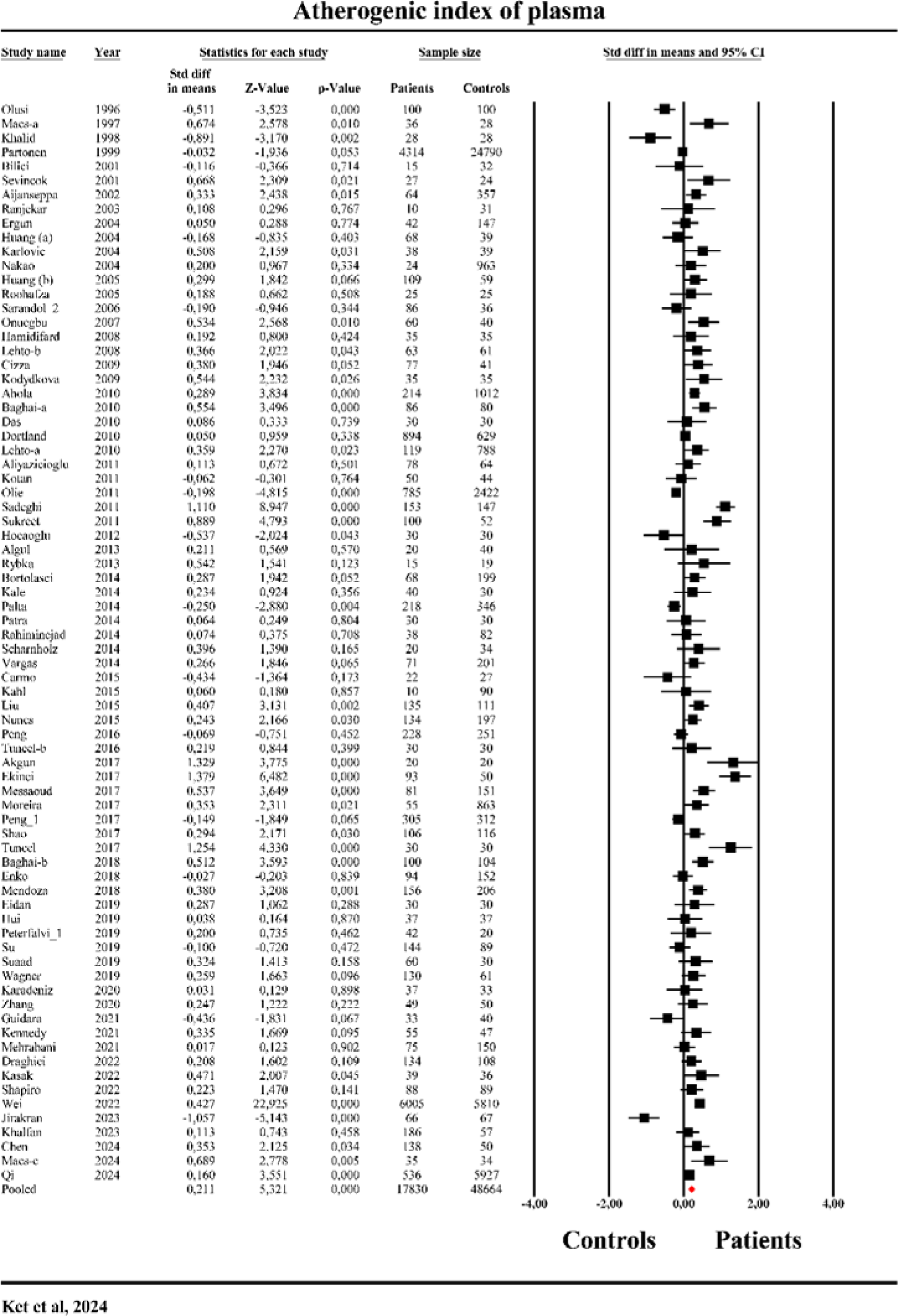
The forest plot of atherogenic index of plasma (AIP) in patients with major depression as compared to healthy controls.

We conducted a meta-analysis of 12 studies to assess the effect size of the ApoB/ApoA ratio. The results showed that two studies had CIs entirely below zero, while four studies had CIs fully above zero. Six studies exhibited overlapping CIs with mixed outcomes: three studies reported negative SMDs, and three reported positive SMDs, as detailed in Table 1. Table 2 and **Figure 4** demonstrate that patients with MDD/BD had significantly higher ApoB/ApoA ratios compared to healthy controls. The publication bias analysis in Table 3 identified three missing studies on the right side of the funnel plot. Including these missing studies would increase the effect size to 0.386.

**Figure 4.**
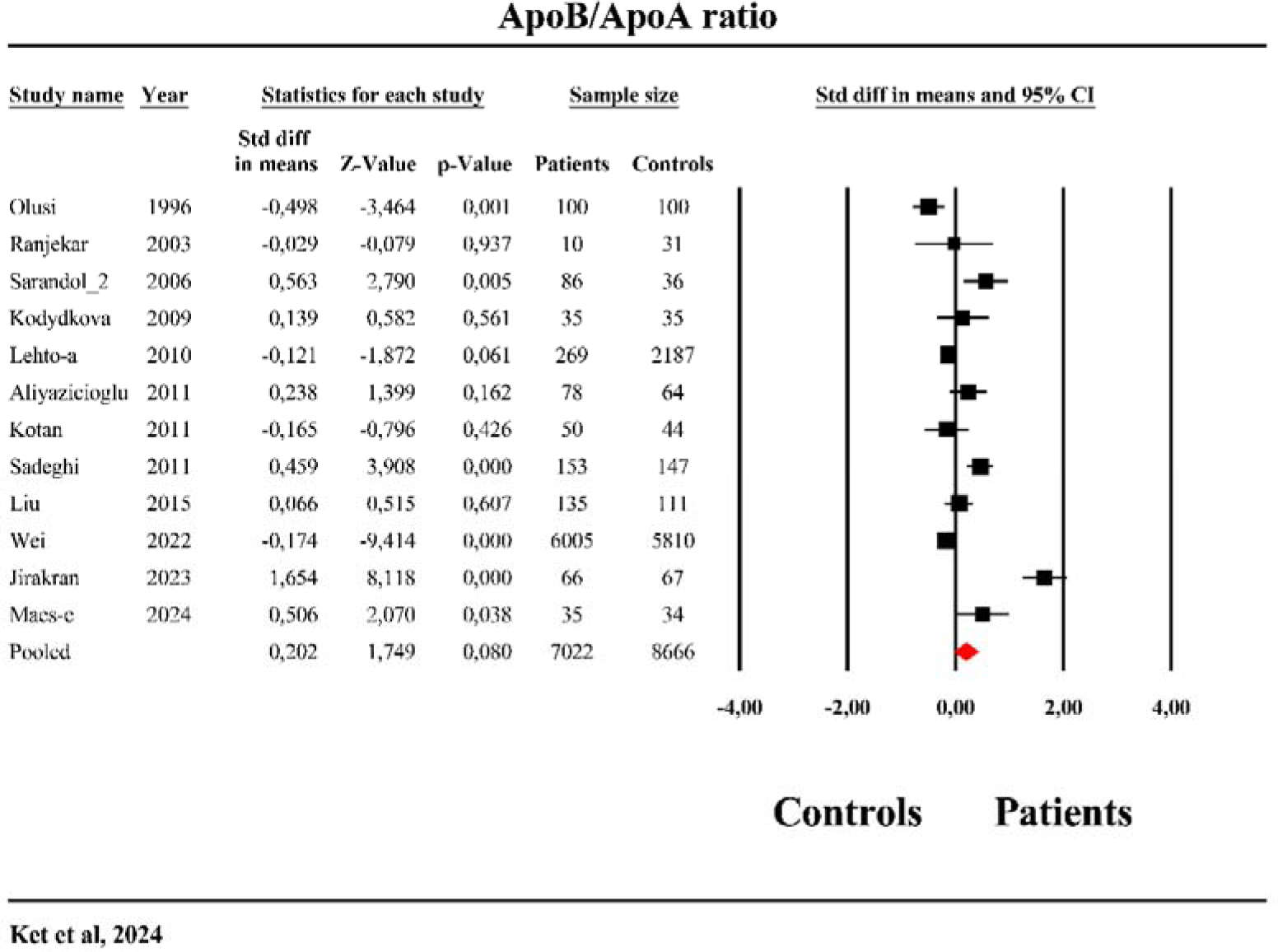
The forest plot of apolipoprotein (Apo)B/ApoA ratio in patients with major depression as compared to healthy controls.

We analyzed data from 78 studies to determine the effect size of the (TG+LDL+VLDL)/(HDL+ApoA) ratio. Our findings show that 5 studies displayed CIs completely below zero, whereas 20 presented CIs above zero. In 53 studies, we found overlapping CIs with varying outcomes; 17 studies documented negative SMD values, and 36 studies reported positive SMD values, as detailed in Table 1. Table 2 and **Figure 5** indicated that the (TG+LDL+VLDL)/(HDL+ApoA) ratio was significantly elevated in patients with MDD/BD compared to healthy controls. Publication bias (Table 3) suggested 12 studies missing from the left side of the funnel plot. Upon imputation, these studies result in a reduction of the SMD to non-significant level.

**Figure 5.**
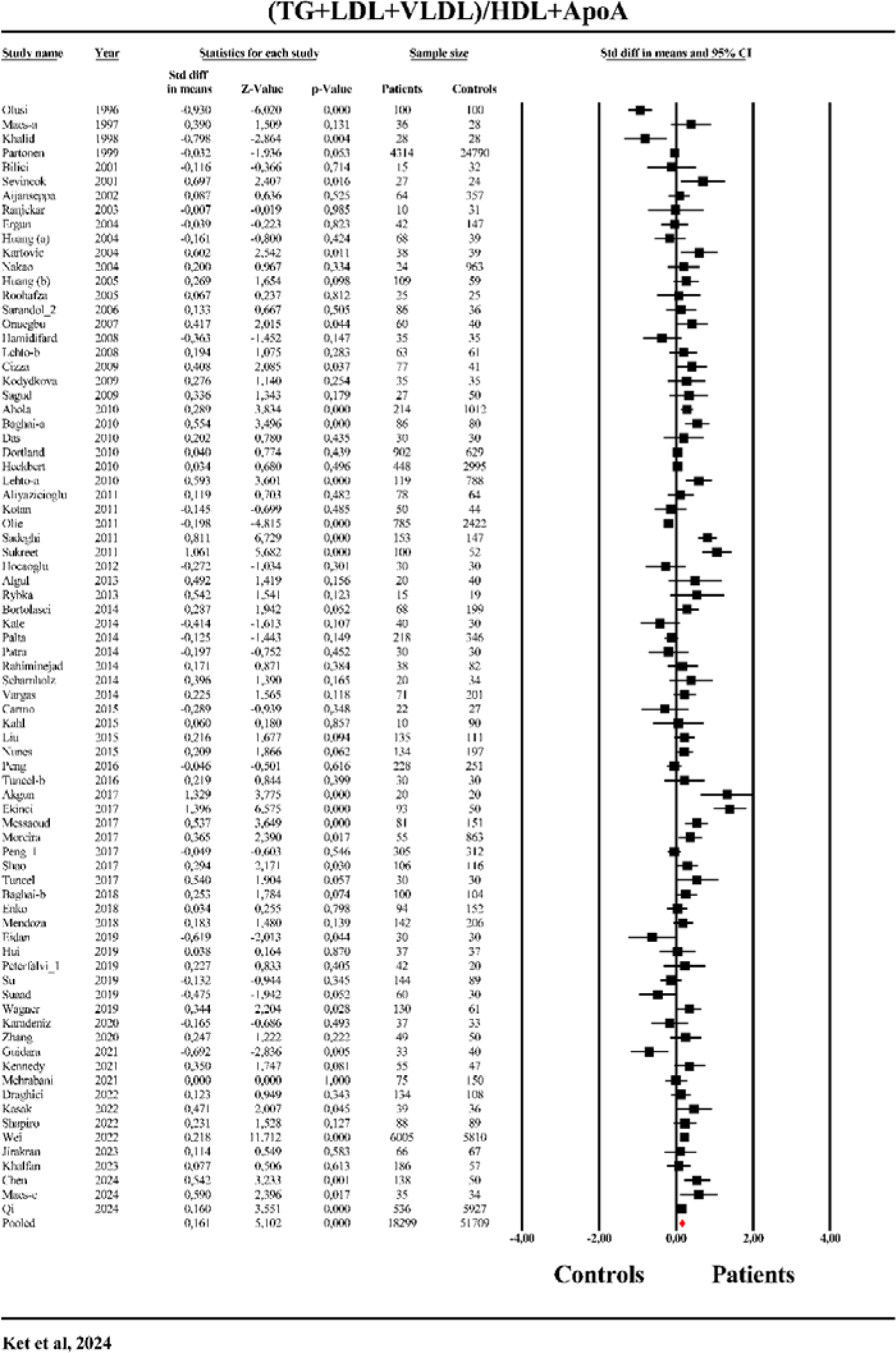
The forest plot of triglyceride (TG)+ low-density lipoprotein (LDL)+very low-density lipoprotein (VLDL)/ high-density lipoprotein (HDL) + Apolipoprotein (ApoA) ratio in patients with major depression as compared to healthy controls.

#### Secondary outcome variables

##### Total cholesterol

This study analyzed data from 75 studies to assess the effect size of total cholesterol. The analysis revealed that 20 studies had CIs entirely below zero, and 13 had CIs exclusively above zero; 42 studies exhibited overlapping CIs with varied outcomes: 19 yielded negative SMD values, and 23 studies reported positive SMD values (see Table 1). The findings presented in Table 2 and ESF, Figure 2 indicate that total cholesterol levels were not significantly different in patients with MDD/BD compared to healthy controls. Egger’s and Kendall’s tests (Table 3) identified publication bias with 12 missing studies on the left side of the funnel plot; incorporating these studies would decrease the effect size rendering a significant inverse association.

##### Triglycerides

We evaluated data from 69 studies to determine the effect size of TG as shown in Table 1. This analysis found that 7 studies had CIs entirely below zero, while 25 studies exhibited CIs above zero. Furthermore, 38 studies showed overlapping CIs with diverse outcomes: 9 reported negative SMD values, and 29 documented positive SMD values. Table 2 and ESF, Figure 3 results indicated that the triglyceride levels were significantly elevated in patients with MDD/BD compared to healthy controls.

##### VLDL

We analyzed data from 9 studies to assess the effect size of the VLDL. As shown in Table 2 and ESF Figure 4, the results indicated no significant difference in VLDL levels between patients with MDD/BD and healthy controls. Additionally, Table 3 did not show evidence of bias in the VLDL findings. The affect size of TG+VLDL was determined by analyzing 69 studies as detailed in Table 1. The results, shown in Table 2 and ESF Figure 5, revealed a significantly elevated TG+VLDL composite score in patients with MDD/BD compared to healthy controls.

##### LDL

We analyzed data from 59 studies to assess the effect size of LDL levels, as shown in Table 1. The findings, displayed in Table 2 and ESF Figure 6, revealed no significant difference in LDL levels between patients with MDD/BD and healthy controls.

##### ApoB

In this study, we assessed ApoB levels using data from 12 studies. As presented in Table 2 and ESF Figure 7, the analysis found no significant differences in ApoB levels between patients with MDD and healthy controls. Table 3 highlights one potentially missing study on the left side of the funnel plot. After including this missing study, there was a slight decrease in the SMD, but the result remained non-significant.

##### HDL

The data from 69 studies was combined to examine the effect size of HDL. Our findings indicated that 25 studies displayed CIs entirely below zero, and 9 showed CIs above zero. A considerable part of the dataset, consisting of 35 studies, presented overlapping CIs with varying results; 23 studies documented negative SMD values, and 12 reported positive SMD values, as detailed in Table 1. HDL levels were significantly lower in MDD/BD patients than in healthy controls, as demonstrated in Table 2 and ESF, Figure 8. Publication bias analysis in Table 3 uncovered 16 missing studies on the left side of the funnel plot. Imputing these studies would further reduce the effect size. Furthermore, group analysis (Table 4) revealed a significant difference (p<0.0001) between studies that included patients with MetS and studies that excluded them.

##### ApoA

We utilized data from 10 studies to assess the effect size of ApoA. The findings in Table 2 and ESF Figure 9 demonstrated no significant difference in ApoA levels between patients with MDD/BD and healthy controls.

##### LCAT enzyme activity

This study evaluated data from 3 studies to determine the effect size of LCAT enzyme activity. The analysis showed that two studies had CIs entirely below zero, while one study had overlapping CIs with a negative SMD. As presented in Table 2 and ESF Figure 10, the results indicate a significant reduction in LCAT enzyme activity in patients with MDD/BD compared to healthy controls. Egger’s and Kendall’s tests, as shown in Table 3, revealed no evidence of bias in the findings.

### Meta-regression analyses

To identify the specific factors contributing to the observed heterogeneity in studies examining atherogenicity in MDD/BD, a meta-regression analysis was conducted. The findings, detailed in ESF Table 6, indicate that the heterogeneity in atherogenicity indices is significantly influenced by the percentage of Caucasian participants, redpoints scores, MetS, sex, and the status of the index of episode.

## Discussion

### Atherogenicity indices and mood disorders

The first significant finding of this meta-analysis is that MDD/BD was substantially associated with two frequently employed atherogenicity indices: the AIP and the Castelli risk index 2, whereas there were no significant associations with Castelli risk index 1 and the ApoB/ApoA ratio. In addition, no differences in any of the indices reported here were found between MDD and BD.

The reason for the absence of an association between MDD/BD and the Castelli risk index 1 can be inferred from the analyses of the individual lipids. Specifically, the numerators of the three atherogenic ratios exhibit variations in their associations with MDD/BD, despite the fact that HDL is the denominator. Thus, triglyceride levels (the numerator in the AIP) were substantially and positively associated with MDD/BD, whereas no such changes were found for LDL (numerator in Castelli risk index 2) and total cholesterol (numerator in Castelli risk index 1). In actuality, the AIP index’s superior performance in comparison to the Castelli risk indices 1 and 2 is due to the distinctions in the numerators, as determined by SMD and z values. As a result, AIP may be perceived as the most critical biomarker for MDD/BD among the classical atherogenicity ratios.

Moreover, the heterogeneity assessments indicate that AIP exhibited the lowest heterogeneity (as measured by the τ2 index) in comparison to both Castelli indices and the ApoB/ApoA index. Additionally, clinical investigations that compared the AIP index with the Castelli risk indices concluded that the AIP is a more effective index for MDD/BD. For instance, Nunes et al. (2015) demonstrated that the AIP and Castelli risk index 2 were elevated in MDD/BD (Nunes, Piccoli de Melo et al. 2015). Conversely, individuals with MDD/BD and concomitant tobacco use disorder (TUD) exhibited elevated atherogenicity levels in comparison to the distinct groups of MDD/BD and TUD patients and controls. Additionally, the association between AIP and MDD/BD was more robust than that of the Castelli risk index 2 with MDD/BD (Nunes, Piccoli de Melo et al. 2015). In a separate investigation, the Castelli risk indices were significantly higher in MDD patients than in controls (Vargas, Nunes et al. 2014). However, another study showed that perhaps the ApoB/ApoA ratio was an adequate biomarker for MDD (Maes, Zhou et al. 2024).

Additionally, the present investigation demonstrated that the heterogeneity of both Castelli risk indices was exacerbated by confounding variables, as evidenced by our “red point score.” This suggests that the heterogeneity of these markers is significantly increased when confounders are not adjusted or controlled for. Most importantly, the presence of MetS significantly impacted both Castelli indices. On the other hand, the heterogeneity in the AIP index did not seem to be influenced by red points or the presence of MetS. The latter is important as this systemic review showed that only 7 out of 85 studies excluded subjects with MetS, whilst the remaining studies did not clearly mention exclusion criteria with regards to MetS. Thus, only 7 papers explicitly stated that they excluded individuals with MetS. This is clearly insufficient, although it suggests that even without accounting for MetS and its components, MDD/BD are associated with heightened atherogenicity, as measured with the AIP index.

In fact, a recent study demonstrated that the examination of the associations between MDD and atherogenicity profiles and separate lipids did not yield any significant results in a combined cohort of MDD and MetS patients (Jirakran, Vasupanrajit et al. 2023). Nevertheless, highly significant associations were observed between atherogenicity indices and MDD after individuals with MetS were excluded. Moreover, even indicants of subclinical MetS (when one or more of the latter 5 MetS components are present) may affect lipid levels measured in MDD (Maes, Vasupanrajit et al. 2024).

Alternatively, we anticipated that a composite of atherogenic compounds in the numerator (e.g., TG + LDL + VLDL) and a composite of antiatherogenic compounds in the denominator (HDL + ApoA) may produce superior outcomes. This ratio was highly significantly associated with MDD/BD and showed lower heterogeneity as compared with the other indices.

Nevertheless, it is also important to investigate publication bias in the different atherogenicity markers. Other pro-arguments to use the AIP are that this index did not seem to be influenced by red points, the presence of MetS, or publication bias. Conversely, substantial publication bias was observed in Castelli risk index 1, the ApoB/ApoA ratio, and the TG + LDL + VLDL / HDL + ApoA ratio. Following the imputation of absent values on the left side, a negative correlation was observed between MDD/BD and the Castelli risk index 1. The ratio of TG + LDL + VLDL to HDL + ApoA lost its significance after imputing missing values, but the ApoB/ApoA ratio gained significance.

All in all, the results of these studies indicate that MDD/BD is associated with an increase in atherogenicity, especially when assessed with the AIP index. The absence of adjustment for MetS or its components may obscure the precise associations with the other atherogenicity indices. Consequently, the detection of any associations is contingent upon the number of individuals with MetS who are included in the study. It appears that the most precise method is to exclude subjects with MetS (Maes et al., 2024).

### Lipid levels in MDD

Our meta-analysis found that MDD and BD were substantially and negatively associated with HDL cholesterol and LCAT, and positively with TG. Nonetheless, the heterogeneity in those individual markers exceeded that observed with the AIP index. The variability of HDL values was more pronounced than that of the AIP and Castelli indices. The primary obstacle is that HDL is significantly affected by the presence of MetS as detected in our group analysis. As with the AIP index, no significant bias was observed in HDL and triglyceride values. These results show that the AIP is a superior index and that its robustness is contingent upon both its numerator and denominator, which are both linked to MDD/BD.

Our systematic evaluation encompassed a limited number of papers on LCAT activity, which indicated an inverse association between LCAT and MDD. Nevertheless, the heterogeneity is large, although no publication bias was present. Consequently, future research should concentrate on the measurements of free cholesterol, cholesteryl esters, and LCAT activity. Instead of LCAT activity based on the assays of free cholesterol (Maes, Delanghe et al. 1994), it is possible to evaluate LCAT concentrations or use a method to directly measure LCAT activity. Nevertheless, the evaluation of LCAT activity derived from free cholesterol and cholesteryl esters is arguably the most appropriate, as it is based on lipid measurements that have functional properties associated with MDD (Maes, Zhou et al. 2024).

### Other confounders

The discovery that there are various MDD/BD subgroups and features that influence atherogenicity is another important confounder that could result in a significant heterogeneity in atherogenicity results. Maes et al. (2024) reported that two subtypes of MDD/BD could be distinguished, namely major dysmood disorder (MDMD) and simple dysmood disorder (SDMD), based on clinical features such as the recurrence of episodes, suicidal behaviors, and the severity of the current episode (Maes, Zhou et al. 2024). The former is the more severe subtype, characterized by a higher recurrence of illness than SDMD, which is the less severe subtype. SDMD is characterized by a low number of episodes or a first episode (Maes, Zhou et al. 2024). The most significant distinctions between the two groups are the immune-inflammatory mediators, T cell activation, growth factors, and atherogenicity profiles (Maes, Rachayon et al. 2022, Maes, Rachayon et al. 2023, Maes, Zhou et al. 2024). In the group of individuals without MetS, it was observed that various atherogenic indices (including ApoB/ApoA) were substantially higher in MDMD than in controls and SDMD (Maes, Zhou et al. 2024). Additionally, the atherogenicity profiles in MDD without MetS were significantly influenced by the recurrence of illness (as measured by the number of episodes and suicidal behaviors) and adverse childhood experiences (Maes, Zhou et al. 2024). The latter study determined that the Research and Diagnostic Algorithmic Rule (RADAR) method, a recently developed diagnostic system, should incorporate ApoB/ApoA and RCT indexes (Maes, Zhou et al. 2024).

It is of the utmost importance to consider and incorporate atherogenicity indices into a diagnostic system for MDD/BD. Firstly, the current body of evidence indicates that there is a strong association between an increased susceptibility to MDD/BD and MetS syndrome or atherogenicity (Nousen, Franco et al. 2013). MetS comorbidity is linked to a more complex affective presentation, a reduced likelihood of recovery, and an increased frequency of episodes and suicide attempts (Fagiolini, Frank et al. 2005, Thomas, Pellicciari et al. 2008, Fries, Pfaffenseller et al. 2012, Grande, Magalhães et al. 2012). Secondly, reduced HDL cholesterol levels in the bloodstream and dysfunctions in the reverse transport of cholesterol can be identified as metabolic risk factors that are shared between mood disorders and CVD, including arteriosclerosis and stroke (Maes, Smith et al. 1997, Maes, Christophe et al. 1999). Third, MDD is characterized by substantial interactions between immune-inflammatory biomarkers and MetS or atherogenicity indices, which result in a more severe phenotype (Maes, Jirakran et al. 2024). In reality, the presence of MetS or atherogenicity in MDD is associated with an aggravation of immune-inflammatory pathways. Additionally, the first episode of SDMD without MetS is distinguished by lowered LCAT activity and increased free cholesterol indicating that this condition is characterized by a pre-atherogenic state even in the absence of MetS (Maes, Vasupanrajit et al. 2024).

### Limitations

The correct evaluation of atherogenicity biomarkers in mood disorders is fraught with numerous sources of heterogeneity, including MetS, race, and the phase of the index episode. Most studies failed to account for MetS or its components. The phenotypic features of mood disorders, such as recurrence of illness and distinct subtypes, may be the most significant confounders, in addition to MetS.

## Conclusions

The findings of this investigation suggest that mood disorders are linked to elevated levels of atherogenicity. The heterogeneity of the data and the presence of bias somewhat impede the selection of the most appropriate atherogenicity biomarker. However, it may seem that the AIP index may be the most effective atherogenic biomarker for MDD/BD. Future research should investigate whether the application of LCAT, free cholesterol, ApoA, or ApoB, measurements can enhance the relevance of atherogenicity biomarkers in the context of MDD.

In any event, the findings of this paper indicate that it is imperative to conduct routine MetS screenings for mood disorder patients. Psychiatrists must consistently assess atherogenicity (by assessing total cholesterol, LDL, HDL, TG, free cholesterol, LCAT activity) and determine the presence of MetS utilizing the American Heart Association criteria (American Heart Association 2023). Patients with MetS or hyperlipidemia should receive treatment for these disorders, since they may exacerbate the phenotype and stage of mood disorders. In the absence of MetS, psychiatrists should assess the AIP as TG/HDL cholesterol. Lipid-lowering therapies (e.g. statins, diets, omega-3 supplements) should be commenced to avert the exacerbation of atherogenicity when the AIP index is increased. It appears that anti-atherogenic therapies are essential to avert recurrence of mood disorders, the exacerbation of the phenome of depression, and suicidal tendencies, all of which are associated with heightened atherogenicity (Maes, Zhou et al. 2024). Similarly, individuals experiencing their first episode of MDD should be assessed for preclinical MetS through the evaluation of LCAT, free cholesterol, and ApoE (Maes, Zhou et al. 2024).

## Author Declarations

### Ethical approval and consent to participate

Not applicable.

### Consent for publication

Not applicable.

### Availability of data and materials

The dataset generated during and/or analyzed during the current study will be available from MM upon reasonable request and once the dataset has been fully exploited by the authors.

## Author’s contributions

The work was designed by MM and KJ. Data were collected by KJ, TJ,AV and PJ. Statistical analyses were performed by MM and AA. All authors revised and approved the final draft.

## Acknowledgements

We are grateful to all participants who participated in this study, nurses, and staff at department of Psychiatry King Chulalongkorn Memorial Hospital, the Thai Red Cross Society, who helped recruit our participants, and Center for Medical Diagnostic Laboratories Faculty of Medicine Chulalongkorn University/King Chulalongkorn Memorial Hospital. This work was supported by the grant from Ratchadapiseksompotch Fund, Graduate Affairs, Faculty of Medicine, Chulalongkorn University (grant numbers RA64/021), Bangkok, Thailand. KJ scholarship from CU Graduate School Thesis Grant and the scholarship from Graduate School, Chulalongkorn University to Commemorate the 72nd anniversary of his Majesty King Bhumibol Adulyadej is gratefully acknowledged.

## References

Ahola, A. J., L. M. Thorn, M. Saraheimo, C. Forsblom, P.-H. Groop and F. S. Group (2010). “Depression is associated with the metabolic syndrome among patients with type 1 diabetes.” Annals of medicine 42(7): 495–501.

Äijänseppä, S., P. Kivinen, E.-L. Helkala, S.-L. Kivelä, J. Tuomilehto and A. Nissinen (2002). “Serum cholesterol and depressive symptoms in elderly Finnish men.” International Journal of Geriatric Psychiatry 17(7): 629–634.

Akgün, S., T. Köken and A. Kahraman (2017). “Evaluation of adiponectin and leptin levels and oxidative stress in bipolar disorder patients with metabolic syndrome treated by valproic acid.” J Psychopharmacol 31(11): 1453–1459.

Al-Amarei, H. M., S. M. H. Rasheed and A. Eidan (2019). “C-reactive protein and its relationship with lipid profile in suicidal and non suicidal adults with major depression.” Indian J Public Health Res Dev 10: 569.

Aliyazicioğlu, R., O. Değer, B. Kural, Ç. Hocaoğlu, M. Colak and F. Yucesan (2011). “The relationship between the peroxisome proliferator-activated receptor gamma 2 gene polymorphism, lipids and adipokines in patients with major depression.” Turkiye Klinikleri Tip Bilimleri Dergisi 31(5).

Almulla, A. F., Y. Thipakorn, A. A. A. Algon, C. Tunvirachaisakul, H. K. Al-Hakeim and M. Maes (2023). “Reverse cholesterol transport and lipid peroxidation biomarkers in major depression and bipolar disorder: A systematic review and meta-analysis.” Brain Behav Immun 113: 374–388.

Almulla, A. F., Y. Thipakorn, A. Vasupanrajit, A. A. Abo Algon, C. Tunvirachaisakul, A. A. Hashim Aljanabi, G. Oxenkrug, H. K. Al-Hakeim and M. Maes (2022). “The tryptophan catabolite or kynurenine pathway in major depressive and bipolar disorder: A systematic review and meta-analysis.” Brain, Behavior, & Immunity - Health 26: 100537.

Almulla, A. F., Y. Thipakorn, A. Vasupanrajit, C. Tunvirachaisakul, G. Oxenkrug, H. K. Al-Hakeim and M. Maes (2022) “The Tryptophan Catabolite or Kynurenine Pathway in a Major Depressive Episode with Melancholia, Psychotic Features and Suicidal Behaviors: A Systematic Review and Meta-Analysis.” Cells 11 DOI: 10.3390/cells11193112.

American Heart Association, A. (2023). “Metabolic Syndrome.” Retrieved 15 Sep, 2024, from https://www.heart.org/en/health-topics/metabolic-syndrome/about-metabolic-syndrome.

Andres-Rodriguez, L., X. Borras, A. Feliu-Soler, A. Perez-Aranda, N. Angarita-Osorio, P. Moreno-Peral, J. Montero-Marin, J. Garcia-Campayo, A. F. Carvalho and M. Maes (2020). “Peripheral immune aberrations in fibromyalgia: A systematic review, meta-analysis and meta-regression.” Brain, behavior, and immunity 87: 881–889.

Baghai, T. C., G. Varallo-Bedarida, C. Born, S. Häfner, C. Schüle, D. Eser, R. Rupprecht, B. Bondy and C. von Schacky (2011). “Major depressive disorder is associated with cardiovascular risk factors and low Omega-3 Index.” J Clin Psychiatry 72(9): 1242–1247.

Baghai, T. C., G. Varallo-Bedarida, C. Born, S. Häfner, C. Schüle, D. Eser, P. Zill, A. Manook, J. Weigl, S. Jooyandeh, C. Nothdurfter, C. von Schacky, B. Bondy and R. Rupprecht (2018). “Classical Risk Factors and Inflammatory Biomarkers: One of the Missing Biological Links between Cardiovascular Disease and Major Depressive Disorder.” Int J Mol Sci 19(6).

Bajwa, W. K., G. M. Asnis, W. C. Sanderson, A. Irfan and H. M. van Praag (1992). “High cholesterol levels in patients with panic disorder.” Am J Psychiatry 149(3): 376–378.

Bilici, M., H. Efe, M. A. Köroğlu, H. A. Uydu, M. Bekaroğlu and O. Değer (2001). “Antioxidative enzyme activities and lipid peroxidation in major depression: alterations by antidepressant treatments.” J Affect Disord 64(1): 43–51.

Bortolasci, C. C., H. O. Vargas, A. Souza-Nogueira, D. S. Barbosa, E. G. Moreira, S. O. V. Nunes, M. Berk, S. Dodd and M. Maes (2014). “Lowered plasma paraoxonase (PON) 1 activity is a trait marker of major depression and PON1 Q192R gene polymorphism–smoking interactions differentially predict the odds of major depression and bipolar disorder.” Journal of affective disorders 159: 23–30.

Chen, Q., T. Sun, Q. He, J. Yu, X. Zhang, L. Han and Y. Ren (2024). “Study of decreased serum levels of C1q/TNF-related protein 4 (CTRP4) in major depressive disorder.” Journal of Psychiatric Research 172: 274–280.

Cizza, G., F. Eskandari, M. Coyle, P. Krishnamurthy, E. C. Wright, S. Mistry and G. Csako (2009). “Plasma CRP levels in premenopausal women with major depression: a 12-month controlled study.” Horm Metab Res 41(8): 641–648.

Cohen, J. (2013). Statistical power analysis for the behavioral sciences, Routledge.

Das, P. P., S. Malhotra, S. Chakrabarti and S. Sharma (2010). “Elevated total cholesterol in severely depressed patients: Role in cardiovascular risk?” The World Journal of Biological Psychiatry 11(2-2): 321–328.

de Melo, L. G. P., S. O. V. Nunes, G. Anderson, H. O. Vargas, D. S. Barbosa, P. Galecki, A. F. Carvalho and M. Maes (2017). “Shared metabolic and immune-inflammatory, oxidative and nitrosative stress pathways in the metabolic syndrome and mood disorders.” Progress in Neuro-Psychopharmacology and Biological Psychiatry 78: 34–50.

Draghici, R. (2022). “The Link between Lipid Profile, Cardiovascular Risk and Mood Disorders Appearance in Older Patients.” Gerontology and Geriatric Medicine 8(3): 1–6.

Eidan, A. J., R. A. Al-Harmoosh and H. M. Al-Amarei (2019). “Estimation of IL-6, INFγ, and Lipid Profile in Suicidal and Nonsuicidal Adults with Major Depressive Disorder.” J Interferon Cytokine Res 39(3): 181–189.

Ekinci, O. and A. Ekinci (2017). “The connections among suicidal behavior, lipid profile and low-grade inflammation in patients with major depressive disorder: a specific relationship with the neutrophil-to-lymphocyte ratio.” Nordic Journal of Psychiatry 71(8): 574–580.

Enko, D., W. Brandmayr, G. Halwachs-Baumann, W. J. Schnedl, A. Meinitzer and G. Kriegshäuser (2018). “Prospective plasma lipid profiling in individuals with and without depression.” Lipids Health Dis 17(1): 149.

Ergün, U. G., S. Uguz, N. Bozdemir, R. Güzel, R. Burgut, E. Saatçi and E. Akpinar (2004). “The relationship between cholesterol levels and depression in the elderly.” Int J Geriatr Psychiatry 19(3): 291–296.

Erman, F., B. Kara, M. Kara and O. Erman (2013). “The Effect of Short-Term Antidepressant Treatment on Serum Levels of Nesfatin-1, Nitric Oxide and Ghrelin in Patients with Major Depressive Disorder.”

Fagiolini, A., E. Frank, J. A. Scott, S. Turkin and D. J. Kupfer (2005). “Metabolic syndrome in bipolar disorder: findings from the Bipolar Disorder Center for Pennsylvanians.” Bipolar Disord 7(5): 424–430.

Fries, G. R., B. Pfaffenseller, L. Stertz, A. V. C. Paz, A. A. Dargél, M. Kunz and F. Kapczinski (2012). “Staging and Neuroprogression in Bipolar Disorder.” Current Psychiatry Reports 14(6): 667–675.

Glomset, J. A., K. R. Norum, A. V. Nichols, W. C. King, C. D. Mitchell, K. R. Applegate, E. L. Gong and E. Gjone (1975). “Plasma Lipoproteins in Familial Lecithin:Cholesterol Acyltransferase Deficiency: Effects of Dietary Manipulation.” Scandinavian Journal of Clinical and Laboratory Investigation 35(sup142): 3–30.

Grande, I., P. V. Magalhães, M. Kunz, E. Vieta and F. Kapczinski (2012). “Mediators of allostasis and systemic toxicity in bipolar disorder.” Physiol Behav 106(1): 46–50.

Guidara, W., M. Messedi, M. Maalej, M. Naifar, W. Khrouf, S. Grayaa, M. Maalej, D. Bonnefont-Rousselot, F. Lamari and F. Ayadi (2021). “Plasma oxysterols: Altered level of plasma 24-hydroxycholesterol in patients with bipolar disorder.” The Journal of Steroid Biochemistry and Molecular Biology 211: 105902.

Hamidifard, S., A. Fakhari, S. Mahboob and B. P. Gargari (2009). “Plasma levels of lipoprotein (a) in patients with major depressive disorders.” Psychiatry Research 169(3): 253–256.

Heckbert, S. R., C. M. Rutter, M. Oliver, L. H. Williams, P. Ciechanowski, E. H. B. Lin, W. J. Katon and M. Von Korff (2010). “Depression in Relation to Long-term Control of Glycemia, Blood Pressure, and Lipids in Patients with Diabetes.” Journal of General Internal Medicine 25(6): 524–529.

Higgins JPT, T. J., Chandler J, Cumpston M, Li T, Page MJ, Welch VA (2019). Cochrane Handbook for Systematic Reviews of Interventions. Chichester (UK), John Wiley & Sons.

Hocaoglu, C., B. Kural, R. Aliyazıcıoglu, O. Deger and S. Cengiz (2012). “IL-1β, IL-6, IL-8, IL-10, IFN-γ, TNF-α and its relationship with lipid parameters in patients with major depression.” Metab Brain Dis 27(4): 425–430.

Huang, T. L. (2005). “Serum lipid profiles in major depression with clinical subtypes, suicide attempts and episodes.” J Affect Disord 86(1): 75–79.

Huang, T. L. and J. F. Chen (2004). “Lipid and lipoprotein levels in depressive disorders with melancholic feature or atypical feature and dysthymia.” Psychiatry Clin Neurosci 58(3): 295–299.

Hui, L., X. L. Yin, J. Chen, X. Y. Yin, H. L. Zhu, J. Li, G. Z. Yin, X. W. Xu, X. N. Yang, Z. K. Qian, C. X. Jiang, Z. Tang, H. B. Yang, E. F. C. Cheung, R. C. K. Chan and Q. F. Jia (2019). “Association between decreased HDL levels and cognitive deficits in patients with bipolar disorder: a pilot study.” Int J Bipolar Disord 7(1): 25.

Jirakran, K., A. Vasupanrajit, C. Tunvirachaisakul, M. Kubera and M. Maes (2023). “Major depression, suicidal behaviors and neuroticism are pro-atherogenic states driven by lowered reverse cholesterol transport.” medRxiv: 2023.2002.2010.23285746.

Jow, G.-M., T.-T. Yang and C.-L. Chen (2006). “Leptin and cholesterol levels are low in major depressive disorder, but high in schizophrenia.” Journal of Affective Disorders 90(1): 21–27.

Kahl, K. G., U. Schweiger, C. Correll, C. Müller, M. L. Busch, M. Bauer and P. Schwarz (2015). “Depression, anxiety disorders, and metabolic syndrome in a population at risk for type 2 diabetes mellitus.” Brain Behav 5(3): e00306.

Kale, A. B., S. B. Kale, S. S. Chalak, R. T. S, G. Bang, M. Agrawal and M. Kaple (2014). “Lipid parameters - significance in patients with endogenous depression.” J Clin Diagn Res 8(1): 17–19.

Karadeniz, S., H. Yaman, Ç. Bilginer, S. Hızarcı Bulut and S. Yaman (2020). “Serum nesfatin-1, ghrelin, and lipid levels in adolescents with first episode drug naïve unipolar depression.” Nord J Psychiatry 74(8): 613–619.

Karlović, D., D. Buljan, M. Martinac and D. Marcinko (2004). “Serum lipid concentrations in Croatian veterans with post-traumatic stress disorder,post-traumatic stress disorder comorbid with major depressive disorder,or major depressive disorder.” J Korean Med Sci 19(3): 431–436.

Kasak, M., M. F. Ceylan, S. T. Hesapcioglu, A. Senat and Ö. Erel (2022). “Peroxisome Proliferator-Activated Receptor Gamma (PPARγ) Levels in Adolescent with Bipolar Disorder and Their Relationship with Metabolic Parameters.” Journal of Molecular Neuroscience 72(6): 1313–1321.

Kennedy, K. G., A. H. Islam, A. Grigorian, L. Fiksenbaum, R. H. B. Mitchell, B. W. McCrindle, B. J. MacIntosh and B. I. Goldstein (2021). “Elevated lipids are associated with reduced regional brain structure in youth with bipolar disorder.” Acta Psychiatr Scand 143(6): 513–525.

Khalfan, A. F., S. C. Campisi, R. F. Lo, B. W. McCrindle and D. J. Korczak (2023). “The association between adolescent depression and dyslipidemia.” Journal of Affective Disorders 338: 239–245.

Khalid, A., N. Lal, J. K. Trivedi, P. K. Dalal, O. P. Asthana, J. S. Srivastava and A. Akhtar (1998). “SERUM LIPIDS : NEW BIOLOGICAL MARKERS IN DEPRESSION ?” Indian Journal of Psychiatry 40(3).

Kim, Y. K. and A. M. Myint (2004). “Clinical application of low serum cholesterol as an indicator for suicide risk in major depression.” J Affect Disord 81(2): 161–166.

Kodydková, J., L. Vávrová, M. Zeman, R. Jirák, J. Macásek, B. Stanková, E. Tvrzická and A. Zák (2009). “Antioxidative enzymes and increased oxidative stress in depressive women.” Clin Biochem 42(13-14): 1368–1374.

Kotan, V. O., E. Sarandol, E. Kirhan, G. Ozkaya and S. Kirli (2011). “Effects of long-term antidepressant treatment on oxidative status in major depressive disorder: a 24-week follow-up study.” Prog Neuropsychopharmacol Biol Psychiatry 35(5): 1284–1290.

Lehto, S. M., J. Hintikka, L. Niskanen, T. Tolmunen, H. Koivumaa-Honkanen, K. Honkalampi and H. Viinamäki (2008). “Low HDL cholesterol associates with major depression in a sample with a 7-year history of depressive symptoms.” Prog Neuropsychopharmacol Biol Psychiatry 32(6): 1557–1561.

Lehto, S. M., L. Niskanen, T. Tolmunen, J. Hintikka, H. Viinamäki, T. Heiskanen, K. Honkalampi, M. Kokkonen and H. Koivumaa-Honkanen (2010). “Low serum HDL-cholesterol levels are associated with long symptom duration in patients with major depressive disorder.” Psychiatry Clin Neurosci 64(3): 279–283.

Liu, X., P. Zheng, X. Zhao, Y. Zhang, C. Hu, J. Li, J. Zhao, J. Zhou, P. Xie and G. Xu (2015). “Discovery and validation of plasma biomarkers for major depressive disorder classification based on liquid chromatography–mass spectrometry.” Journal of proteome research 14(5): 2322–2330.

Maes, M., A. Christophe, J. Delanghe, C. Altamura, H. Neels and H. Y. Meltzer (1999). “Lowered ω3 polyunsaturated fatty acids in serum phospholipids and cholesteryl esters of depressed patients.” Psychiatry Research 85(3): 275–291.

Maes, M., J. Delanghe, H. Y. Meltzer, S. Scharpé, P. D’Hondt and P. Cosyns (1994). “Lower degree of esterification of serum cholesterol in depression: relevance for depression and suicide research.” Acta Psychiatr Scand 90(4): 252–258.

Maes, M., K. Jirakran, A. Vasupanrajit, B. Zhou, C. Tunvirachaisakul, D. St. Stoyanov and A. F. Almulla (2024). “Are abnormalities in lipid metabolism, together with adverse childhood experiences, the silent causes of immune-linked neurotoxicity in major depression?” medRxiv: 2024.2002.2020.24302841.

Maes, M., M. Kubera, E. Obuchowiczwa, L. Goehler and J. Brzeszcz (2011). “Depression’s multiple comorbidities explained by (neuro)inflammatory and oxidative & nitrosative stress pathways.” Neuro Endocrinol Lett 32(1): 7–24.

Maes, M., M. Rachayon, K. Jirakran, P. Sodsai, S. Klinchanhom, P. Gałecki, A. Sughondhabirom and A. Basta-Kaim (2022). “The Immune Profile of Major Dysmood Disorder: Proof of Concept and Mechanism Using the Precision Nomothetic Psychiatry Approach.” Cells 11(7).

Maes, M., M. Rachayon, K. Jirakran, P. Sodsai and A. Sughondhabirom (2023). “Lower Nerve Growth Factor Levels in Major Depression and Suicidal Behaviors: Effects of Adverse Childhood Experiences and Recurrence of Illness.” Brain Sci 13(7).

Maes, M., P. Ruckoanich, Y. S. Chang, N. Mahanonda and M. Berk (2011). “Multiple aberrations in shared inflammatory and oxidative & nitrosative stress (IO&NS) pathways explain the co-association of depression and cardiovascular disorder (CVD), and the increased risk for CVD and due mortality in depressed patients.” Prog Neuropsychopharmacol Biol Psychiatry 35(3): 769–783.

Maes, M., R. Smith, A. Christophe, E. Vandoolaeghe, A. Van Gastel, H. Neels, P. Demedts, A. Wauters and H. Y. Meltzer (1997). “Lower serum high-density lipoprotein cholesterol (HDL-C) in major depression and in depressed men with serious suicidal attempts: relationship with immune-inflammatory markers.” Acta Psychiatr Scand 95(3): 212–221.

Maes, M., A. Vasupanrajit, K. Jirakran, B. Zhou, C. Tunvirachaisakul and A. F. Almulla (2024). “First-episode mild depression in young adults is a pre-proatherogenic condition even in the absence of subclinical metabolic syndrome: lowered lecithin-cholesterol acyltransferase as a key factor.” medRxiv: 2024.2001.2001.24300688.

Maes, M., B. Zhou, K. Jirakran, A. Vasupanrajit, P. Boonchaya-Anant, C. Tunvirachaisakul, X. Tang, J. Li and A. F. Almulla (2024). “Towards a major methodological shift in depression research by assessing continuous scores of recurrence of illness, lifetime and current suicidal behaviors and phenome features.” J Affect Disord 350: 728–740.

Messaoud, A., R. Mensi, A. Mrad, A. Mhalla, I. Azizi, B. Amemou, I. Trabelsi, M. H. Grissa, N. H. Salem, A. Chadly, W. Douki, M. F. Najjar and L. Gaha (2017). “Is low total cholesterol levels associated with suicide attempt in depressive patients?” Ann Gen Psychiatry 16: 20.

Moreira, F. P., K. Jansen, T. d. A. Cardoso, T. C. Mondin, P. V. d. S. Magalhães, F. Kapczinski, L. D. d. M. Souza, R. A. da Silva, J. P. Oses and C. D. Wiener (2017). “Metabolic syndrome in subjects with bipolar disorder and major depressive disorder in a current depressive episode: Population-based study: Metabolic syndrome in current depressive episode.” Journal of Psychiatric Research 92: 119–123.

Nakao, M. and E. Yano (2004). “Relationship between major depression and high serum cholesterol in Japanese men.” The Tohoku journal of experimental medicine 204(4): 273–287.

Nousen, E. K., J. G. Franco and E. L. Sullivan (2013). “Unraveling the mechanisms responsible for the comorbidity between metabolic syndrome and mental health disorders.” Neuroendocrinology 98(4): 254–266.

Nunes, S. O. V., L. G. Piccoli de Melo, M. R. Pizzo de Castro, D. S. Barbosa, H. O. Vargas, M. Berk and M. Maes (2015). “Atherogenic index of plasma and atherogenic coefficient are increased in major depression and bipolar disorder, especially when comorbid with tobacco use disorder.” Journal of Affective Disorders 172: 55–62.

Olié, E., M. C. Picot, S. Guillaume, M. Abbar and P. Courtet (2011). “Measurement of total serum cholesterol in the evaluation of suicidal risk.” Journal of Affective Disorders 133(1): 234–238.

Olusi, S. O. and A. A. Fido (1996). “Serum lipid concentrations in patients with major depressive disorder.” Biol Psychiatry 40(11): 1128–1131.

Onuegbu, A., E. Agbedana, O. Baiyewu, M. Olisekodiaka, M. Ebesunun, K. Adebayo, O. Ayelagbe and O. Adegoke (2007). “Evaluation of plasma lipids and lipoproteins in Nigerians suffering from depressive illness.” African Journal of Biomedical Research 10(2).

Ormonde do Carmo, M. B., A. C. Mendes-Ribeiro, C. Matsuura, V. L. Pinto, W. V. Mury, N. O. Pinto, M. B. Moss, M. R. Ferraz and T. M. Brunini (2015). “Major depression induces oxidative stress and platelet hyperaggregability.” J Psychiatr Res 61: 19–24.

Oxenkrug, G. F., R. J. Branconnier, N. Harto-Truax and J. O. Cole (1983). “Is serum cholesterol a biological marker for major depressive disorder?” Am J Psychiatry 140(7): 920–921.

Page, M. J., J. E. McKenzie, P. M. Bossuyt, I. Boutron, T. C. Hoffmann, C. D. Mulrow, L. Shamseer, J. M. Tetzlaff, E. A. Akl, S. E. Brennan, R. Chou, J. Glanville, J. M. Grimshaw, A. Hróbjartsson, M. M. Lalu, T. Li, E. W. Loder, E. Mayo-Wilson, S. McDonald, L. A. McGuinness, L. A. Stewart, J. Thomas, A. C. Tricco, V. A. Welch, P. Whiting and D. Moher (2021). “The PRISMA 2020 statement: An updated guideline for reporting systematic reviews.” PLOS Medicine 18(3): e1003583.

Palta, P., S. H. Golden, J. A. Teresi, W. Palmas, P. Trief, R. S. Weinstock, S. Shea, J. J. Manly and J. A. Luchsinger (2014). “Depression is not associated with diabetes control in minority elderly.” Journal of Diabetes and its Complications 28(6): 798–804.

Partonen, T., J. Haukka, J. Virtamo, P. Taylor and J. Lönnqvist (1999). “Association of low serum total cholesterol with major depression and suicide.” The British Journal of Psychiatry 175(3): 259–262.

Patra, B. N., S. K. Khandelwal, R. K. Chadda and L. Ramakrishnan (2014). “A controlled study of serum lipid profiles in Indian patients with depressive episode.” Indian J Psychol Med 36(2): 129–133.

Peng, Y.-F., Y. Xiang and Y.-S. Wei (2016). “The significance of routine biochemical markers in patients with major depressive disorder.” Scientific Reports 6(1): 34402.

Peng, Y. F., S. M. Zhong and Y. H. Qin (2017). “The relationship between major depressive disorder and glucose parameters: A cross-sectional study in a Chinese population.” Adv Clin Exp Med 26(4): 665–669.

Péterfalvi, Á., N. Németh, R. Herczeg, T. Tényi, A. Miseta, B. Czéh and M. Simon (2019). “Examining the Influence of Early Life Stress on Serum Lipid Profiles and Cognitive Functioning in Depressed Patients.” Frontiers in Psychology 10.

Qi, X., S. Wang, Q. Huang, X. Chen, L. Qiu, K. Ouyang and Y. Chen (2024). “The association between non-high-density lipoprotein cholesterol to high-density lipoprotein cholesterol ratio (NHHR) and risk of depression among US adults: A cross-sectional NHANES study.” Journal of Affective Disorders 344: 451–457.

Rahiminejad, M. E., A. Moaddab, S. Rabiee, F. Esna-Ashari, S. Borzouei and S. M. Hosseini (2014). “The relationship between clinicobiochemical markers and depression in women with polycystic ovary syndrome.” Iran J Reprod Med 12(12): 811–816.

Ranjekar, P. K., A. Hinge, M. V. Hegde, M. Ghate, A. Kale, S. Sitasawad, U. V. Wagh, V. B. Debsikdar and S. P. Mahadik (2003). “Decreased antioxidant enzymes and membrane essential polyunsaturated fatty acids in schizophrenic and bipolar mood disorder patients.” Psychiatry Res 121(2): 109–122.

Roohafza, H., M. Sadeghi, H. Afshar, G. Mousavi and S. Shirani (2010). “EVALUATION OF LIPID PROFILE IN PATIENT WITH MAJOR DEPRESSIVE DISORDER AND GENERALIZED ANXIETY DISORDER.” ARYA Atherosclerosis Journal 1(1).

Ruljancic, N., M. Mihanovic and I. Cepelak (2011). “Thrombocyte serotonin and serum cholesterol concentration in suicidal and non-suicidal depressed patients.” Progress in Neuro-Psychopharmacology and Biological Psychiatry 35(5): 1261–1267.

Rybka, J., K. Kędziora-Kornatowska, P. Banaś-Leżańska, I. Majsterek, L. A. Carvalho, A. Cattaneo, C. Anacker and J. Kędziora (2013). “Interplay between the pro-oxidant and antioxidant systems and proinflammatory cytokine levels, in relation to iron metabolism and the erythron in depression.” Free Radical Biology and Medicine 63: 187–194.

Sadeghi, M., H. Roohafza, H. Afshar, F. Rajabi, M. Ramzani, H. Shemirani and N. Sarafzadeghan (2011). “Relationship between depression and apolipoproteins A and B: a case-control study.” Clinics (Sao Paulo) 66(1): 113–117.

Sagud, M., A. Mihaljevic-Peles, N. Pivac, M. Jakovljevic and D. Muck-Seler (2009). “Lipid levels in female patients with affective disorders.” Psychiatry Res 168(3): 218–221.

Sanyal, D., I. Chakrabarti and J. Basu (2000). “Serum cholesterol as a marker in panic disorder: a pilot study.” INTERNATIONAL MEDICAL JOURNAL-TOKYO-7(4): 277–282.

Sarandol, A., E. Sarandol, S. S. Eker, E. U. Karaagac, B. Z. Hizli, M. Dirican and S. Kirli (2006). “Oxidation of apolipoprotein B-containing lipoproteins and serum paraoxonase/arylesterase activities in major depressive disorder.” Prog Neuropsychopharmacol Biol Psychiatry 30(6): 1103–1108.

Scharnholz, B., M. Gilles, A. Marzina, M. Kommer, F. Lederbogen, S. A. Wudy, M. F. Hartmann, S. Westphal, H. J. Roth, K. G. Kahl, A. Meyer-Lindenberg, H. J. Michaely and M. Deuschle (2014). “Do depressed patients without activation of the hypothalamus–pituitary–adrenal (HPA) system have metabolic disturbances?” Psychoneuroendocrinology 39: 104–110.

Segoviano-Mendoza, M., M. Cárdenas-de la Cruz, J. Salas-Pacheco, F. Vázquez-Alaniz, O. La Llave-León, F. Castellanos-Juárez, J. Méndez-Hernández, M. Barraza-Salas, E. Miranda-Morales, O. Arias-Carrión and E. Méndez-Hernández (2018). “Hypocholesterolemia is an independent risk factor for depression disorder and suicide attempt in Northern Mexican population.” BMC Psychiatry 18(1): 7.

Sevincok, L., A. Buyukozturk and F. Dereboy (2001). “Serum lipid concentrations in patients with comorbid generalized anxiety disorder and major depressive disorder.” Can J Psychiatry 46(1): 68–71.

Shapiro, L. R., K. G. Kennedy, M. K. Dimick and B. I. Goldstein (2022). “Elevated atherogenic lipid profile in youth with bipolar disorder during euthymia and hypomanic/mixed but not depressive states.” J Psychosom Res 156: 110763.

Sonal Sukreet, M. B., Mahima B. Subramanyam, Madhur Agrawal, Abhishek and J. K. S. Chaturvedi, Devaramane Virupaksha, Panambur V. Bhandary, Mungli Prakash (2011). “Serum malondialdehyde and thiol levels in patients with bipolar disorder.” BioChemistry: An Indian Journal 1(5).

Su, M., E. Li, C. Tang, Y. Zhao, R. Liu and K. Gao (2019). “Comparison of blood lipid profile/thyroid function markers between unipolar and bipolar depressed patients and in depressed patients with anhedonia or suicidal thoughts.” Mol Med 25(1): 51.

Thomas, C., R. Pellicciari, M. Pruzanski, J. Auwerx and K. Schoonjans (2008). “Targeting bile-acid signalling for metabolic diseases.” Nat Rev Drug Discov 7(8): 678–693.

Tunçel Ö, K., S. Akbaş and B. Bilgici (2016). “Increased Ghrelin Levels and Unchanged Adipocytokine Levels in Major Depressive Disorder.” J Child Adolesc Psychopharmacol 26(8): 733–739.

Tunçel Ö, K., G. Sarısoy, B. Bilgici, O. Pazvantoğlu, E. Çetin and E. K. Tunçel (2018). “Adipocytokines and ghrelin level of bipolar patients from manic episode to euthymic episode.” Nord J Psychiatry 72(2): 150–156.

Vaghef-Mehrabani, E., A. Izadi and M. Ebrahimi-Mameghani (2021). “The association of depression with metabolic syndrome parameters and malondialdehyde (MDA) in obese women: A case-control study.” Health Promot Perspect 11(4): 492–497.

van Reedt Dortland, A. K., E. J. Giltay, T. van Veen, J. van Pelt, F. G. Zitman and B. W. Penninx (2010). “Associations between serum lipids and major depressive disorder: results from the Netherlands Study of Depression and Anxiety (NESDA).” J Clin Psychiatry 71(6): 729–736.

Vargas, H. O., S. O. V. Nunes, D. S. Barbosa, M. M. Vargas, A. Cestari, S. Dodd, K. Venugopal, M. Maes and M. Berk (2014). “Castelli risk indexes 1 and 2 are higher in major depression but other characteristics of the metabolic syndrome are not specific to mood disorders.” Life Sciences 102(1): 65–71.

Vasupanrajit, A., K. Jirakran, C. Tunvirachaisakul, M. Solmi and M. Maes (2022). “Inflammation and nitro-oxidative stress in current suicidal attempts and current suicidal ideation: a systematic review and meta-analysis.” Molecular Psychiatry 27(3): 1350–1361.

Wagner, C. J., C. Musenbichler, L. Böhm, K. Färber, A.-I. Fischer, F. von Nippold, M. Winkelmann, T. Richter-Schmidinger, C. Mühle, J. Kornhuber and B. Lenz (2019). “LDL cholesterol relates to depression, its severity, and the prospective course.” Progress in Neuro-Psychopharmacology and Biological Psychiatry 92: 405–411.

Walker, E. R., R. E. McGee and B. G. Druss (2015). “Mortality in mental disorders and global disease burden implications: a systematic review and meta-analysis.” JAMA Psychiatry 72(4): 334–341.

Wan, X., W. Wang, J. Liu and T. Tong (2014). “Estimating the sample mean and standard deviation from the sample size, median, range and/or interquartile range.” BMC medical research methodology 14(1): 1–13.

Wei, Y., T. Wang, G. Li, J. Feng, L. Deng, H. Xu, L. Yin, J. Ma, D. Chen and J. Chen (2022). “Investigation of systemic immune-inflammation index, neutrophil/high-density lipoprotein ratio, lymphocyte/high-density lipoprotein ratio, and monocyte/high-density lipoprotein ratio as indicators of inflammation in patients with schizophrenia and bipolar disorder.” Frontiers in Psychiatry 13.

Zhang, C., Y. Yang, D.-m. Zhu, W. Zhao, Y. Zhang, B. Zhang, Y. Wang, J. Zhu and Y. Yu (2020). “Neural correlates of the association between depression and high density lipoprotein cholesterol change.” Journal of Psychiatric Research 130: 9–18.

