## supplementary file for "Increased atherogenicity in mood disorders: a systematic review, meta-analysis and meta-regression"

**Biomarkers of metabolic syndrome in major depression: A systematic review, meta-analysis and meta-regression**

Running title: Major depression and metabolic syndrome

**ESF, Table 1. Search sentences and terms used in each database**

| **Database Name** | **Search Sentence** | **No. of Articles** |
| --- | --- | --- |
| **PubMed/Medline** | ((((((((((PON 1 and Depression) OR (PON 1 and MDD)) OR (PON 1 and Mood disorder))OR (depression and paraoxonase 1)) OR (bipolar disorder and paraoxonase 1)) OR (Depression and Apo A)) OR (Depression and Apo B). | **234** |
|  | (HDL and MDD) OR (HDL and Depression)) OR (HDL and Bipolar disorder)) OR (Lipid antioxidants and MDD)) OR (MDD and PON 1)) OR (Bipolar disorder and PON 1)) OR (MDD and oxidized LDL)) OR (MDD and Apo A)) OR (MDD and LDL)) OR (MDD and TG)) OR (MDD and Apo B). | **1,544** |
|  | (HDL and MDD) OR (HDL and Depression)) OR (HDL and Bipolar disorder)) OR (Lipid antioxidants and HDL)) OR (HDL and PON 1)) OR (HDL and PON 1)) OR (HDL and oxidized LDL)) OR (HDL and Apo A)) OR (HDL and LDL)) OR (HDL and TG)) OR (HDL and Apo B). | **64,784** |
|  | (oxidized LDL and MDD) OR (oxidized LDL and Depression)) OR (oxidized LDL and Bipolar disorder)) OR (Lipid antioxidants and MDD)) OR (MDD and PON 1)) OR (Bipolar disorder and PON 1)) OR (MDD and oxidized LDL)) OR (MDD and Apo A)) OR (MDD and LDL)) OR (MDD and TG)) OR (MDD and Apo B) | **267** |
|  | (oxidized LDL and MDD) OR (oxidized LDL and Depression)) OR (oxidized LDL and Bipolar disorder)) OR (Lipid antioxidants and MDD)) OR (mda and pon 1)) OR (Bipolar disorder and PON 1)) OR (mda and oxidized ldl)) OR (mda and apo a)) OR (mda and ldl)) OR (mda and tg)) OR (mda and apo b). | **3,403** |
|  | (Castelli 1 and MDD) OR (Castelli 1 and Depression)) OR (Castelli 1 and Bipolar disorder)) OR (Castelli 1 and PON 1)) OR (Bipolar disorder and PON 1)) OR (MDD and oxidized LDL)) OR (Castelli 1 and Apo A)) OR (Castelli 1 and LDL)) OR (Castelli 1 and TG)) OR (Castelli 1 and Apo B) | **156** |
| **Google Scholar** | ((((((((((PON 1 and Depression) OR (PON 1 and MDD)) OR (PON 1 and Mood disorder))OR (depression and paraoxonase 1)) OR (bipolar disorder and paraoxonase 1)) OR (Depression and Apo A)) OR (Depression and Apo B). | **238** |
|  | (HDL and MDD) OR (HDL and Depression)) OR (HDL and Bipolar disorder)) OR (Lipid antioxidants and MDD)) OR (MDD and PON 1)) OR (Bipolar disorder and PON 1)) OR (MDD and oxidized LDL)) OR (MDD and Apo A)) OR (MDD and LDL)) OR (MDD and TG)) OR (MDD and Apo B). | **4,570** |
|  | (HDL and MDD) OR (HDL and Depression)) OR (HDL and Bipolar disorder)) OR (Lipid antioxidants and HDL)) OR (HDL and PON 1)) OR (HDL and PON 1)) OR (HDL and oxidized LDL)) OR (HDL and Apo A)) OR (HDL and LDL)) OR (HDL and TG)) OR (HDL and Apo B). | **109** |
|  | (oxidized LDL and MDD) OR (oxidized LDL and Depression)) OR (oxidized LDL and Bipolar disorder)) OR (Lipid antioxidants and MDD)) OR (MDD and oxidized LDL)) OR (MDD and Apo A)) OR (MDD and LDL)) OR (MDD and TG). | **693** |
|  | (Castelli 1 and MDD) OR (Castelli 1 and Depression)) OR (Castelli 1 and Bipolar disorder)) OR (Castelli 1 and PON 1)) OR (Bipolar disorder and PON 1)) OR (MDD and oxidized LDL)) OR (Castelli 1 and Apo A)) OR (Castelli 1 and LDL)) OR (Castelli 1 and TG)) OR (Castelli 1 and Apo B) | **575** |

**ESF, Table 2. Immune cofounder’s scale (ICS) applied from Andrés-Rodríguez, et al., 2019**

| **Methodological quality of the study** | |
| --- | --- |
| **1** | Study sample ≥ 128 participants including patients and controls (1= Yes, 0 = No) |
| **2** | Did the study control the results for potential confounders (e.g., age, BMI, gender, race)? (1= Yes, 0 = No) |
| **3** | Were participants with MDD and controls age- and-gender-matched or was there a statistical control? (1= Yes, 0 = No) |
| **4** | Was the time of sample collection specified (e.g., morning vs. evening)? (1= Yes, 0 = No) |
| **5** | Were participants with MDD free of immunomodulatory drugs including anti-cytokines, glucocorticoids, immunoglobulins, and immunosuppressants, or was there a medication washout period or was drug intake statistically controlled for? (1= Yes, 0 = No) |
| **6** | Were participants with affective disorders free of antistatistically controlled for? (1= Yes, 0 = No) |
| **7** | Reporting either the manufacturer of the test or detection limit and coefficients of variation (1= Yes, 0 = No) |
| **8** | Reporting blood fraction (serum, plasma, culture supernatant or whole blood) (1= Yes, 0 = No) |
| **Total quality score (8 points)** | |
| **Biomarker confounders red points**  *The red points should not be given if the item is statistically controlled for* | |
| **1** | 3 red points for comorbid illnesses such as autoimmune disorders & other immune disorders including rheumatoid arthritis, psoriasis, inflammatory bowel disease, chronic obstructive pulmonary disease, multiple sclerosis |
| **2** | 3 red points for use of recreational drugs such as methamphetamine or opioids |
| **3** | 2 red points when groups were not matched for age |
| **4** | 2 red points when groups were not matched for sex |
| **5** | 2 red points for medication use as for example immunomodulators |
| **6** | 2 red points for early traumatic life events |
| **7** | 2 red points for shift work and primary sleep disorders |
| **8** | 1.5 red points for use of antidepressants |
| **9** | 1 red point for other neuro-psychiatric comorbidities, as for example schizophrenia, autism, GAD, PTSD |
| **10** | 1 red point for not fasting (8 hours before blood extraction) |
| **11** | 1 red point for use of omega-3 and antioxidant supplements |
| **12** | 1 red point when data were not controlled for body mass index |
| **13** | 1 red point when data were not controlled for physical activity or sedentary life |
| **14** | 1 red point when data were not controlled for smoking |
| **15** | 1 red point for use of oral contraceptives or NSAIDs |
| **16** | 0.5 red points when data were not controlled for ethnicity in countries such as US, Brazil |
| **17** | 0.5 red points when data were not controlled for seasonality |
| **18** | 0.5 red points when data were not controlled for diurnal variation (8-10 a.m. versus all other time points) |
|  | **Total red point score (26 points)** |

**ESF, Table 3. PRISMA checklist**

| **Section/topic** | **#** | **Checklist item** | **Reported on page #** |
| --- | --- | --- | --- |
| **TITLE** | | | |
| Title | 1 | Identify the report as a systematic review, meta-analysis, or both. | 1 |
| **ABSTRACT** | | | |
| Structured summary | 2 | Provide a structured summary including, as applicable: background; objectives; data sources; study eligibility criteria, participants, and interventions; study appraisal and synthesis methods; results; limitations; conclusions and implications of key findings; systematic review registration number. | 3 |
| **INTRODUCTION** | | | |
| Rationale | 3 | Describe the rationale for the review in the context of what is already known. |  |
| Objectives | 4 | Provide an explicit statement of questions being addressed with reference to participants, interventions, comparisons, outcomes, and study design (PICOS). |  |
| **METHODS** | | | |
| Protocol and registration | 5 | Indicate if a review protocol exists, if and where it can be accessed (e.g., Web address), and, if available, provide registration information including registration number. |  |
| Eligibility criteria | 6 | Specify study characteristics (e.g., PICOS, length of follow-up) and report characteristics (e.g., years considered, language, publication status) used as criteria for eligibility, giving rationale. |  |
| Information sources | 7 | Describe all information sources (e.g., databases with dates of coverage, contact with study authors to identify additional studies) in the search and date last searched. |  |
| Search | 8 | Present full electronic search strategy for at least one database, including any limits used, such that it could be repeated. |  |
| Study selection | 9 | State the process for selecting studies (i.e., screening, eligibility, included in systematic review, and, if applicable, included in the meta-analysis). |  |
| Data collection process | 10 | Describe method of data extraction from reports (e.g., piloted forms, independently, in duplicate) and any processes for obtaining and confirming data from investigators. |  |
| Data items | 11 | List and define all variables for which data were sought (e.g., PICOS, funding sources) and any assumptions and simplifications made. | ESF, Table 1 |
| Risk of bias in individual studies | 12 | Describe methods used for assessing risk of bias of individual studies (including specification of whether this was done at the study or outcome level), and how this information is to be used in any data synthesis. | Table 3 |
| Summary measures | 13 | State the principal summary measures (e.g., risk ratio, difference in means). |  |
| Synthesis of results | 14 | Describe the methods of handling data and combining results of studies, if done, including measures of consistency (e.g., I^2^) for each meta-analysis. |  |
| Risk of bias across studies | 15 | Specify any assessment of risk of bias that may affect the cumulative evidence (e.g., publication bias, selective reporting within studies). | Table 3 |
| Additional analyses | 16 | Describe methods of additional analyses (e.g., sensitivity or subgroup analyses, meta-regression), if done, indicating which were pre-specified. | ESF, Table 9 |
| **RESULTS** | | |  |
| Study selection | 17 | Give numbers of studies screened, assessed for eligibility, and included in the review, with reasons for exclusions at each stage, ideally with a flow diagram. |  |
| Study characteristics | 18 | For each study, present characteristics for which data were extracted (e.g., study size, PICOS, follow-up period) and provide the citations. | ESF, Table 5 |
| Risk of bias within studies | 19 | Present data on risk of bias of each study and, if available, any outcome level assessment (see item 12). | Table 3 |
| Results of individual studies | 20 | For all outcomes considered (benefits or harms), present, for each study: (a) simple summary data for each intervention group (b) effect estimates and confidence intervals, ideally with a forest plot. | Table 1 |
| Synthesis of results | 21 | Present results of each meta-analysis done, including confidence intervals and measures of consistency. | Table 2 |
| Risk of bias across studies | 22 | Present results of any assessment of risk of bias across studies (see Item 15). | Table 3 |
| Additional analysis | 23 | Give results of additional analyses, if done (e.g., sensitivity or subgroup analyses, meta-regression [see Item 16]). |  |
| **DISCUSSION** | | |  |
| Summary of evidence | 24 | Summarize the main findings including the strength of evidence for each main outcome; consider their relevance to key groups (e.g., healthcare providers, users, and policy makers). |  |
| Limitations | 25 | Discuss limitations at study and outcome level (e.g., risk of bias), and at review-level (e.g., incomplete retrieval of identified research, reporting bias). |  |
| Conclusions | 26 | Provide a general interpretation of the results in the context of other evidence, and implications for future research. |  |
| **FUNDING** | | |  |
| Funding | 27 | Describe sources of funding for the systematic review and other support (e.g., supply of data), role of funders for the systematic review. |  |

**EFS, Table 4.** **Studies excluded from the meta-analysis but included in the systematic review**.

| **Authors, year** | **Reason why excluded from the meta-analysis** |
| --- | --- |
| Almeida-Montes et al. 2000 | The authors did not show the mean and standard deviation of the measured lipid biomarkers |
| Baek et al. 2014 | The authors did not show the mean and standard deviation of the measured lipid biomarkers |
| Banikazemi et al. 2015 | The authors did not show the mean and standard deviation of the measured lipid biomarkers |
| Park et al. 2013 | The authors did not show the mean and standard deviation of the measured lipid biomarkers |
| Sullivan et al. 1994 | The authors did not show the mean and standard deviation of the measured lipid biomarkers |
| Xu et al. 2024 | The authors did not show the mean and standard deviation of the measured lipid biomarkers |

**ESF, Table 5. Characteristics of studies which were included in the systematic reviews and meta-analysis**

| **#** | **Authors, years** | **Setting** | **Type of case** | **Type of Control** | **Sample Size** | | | | **Age** | | **Assessed**  **Biomarkers** | **Specimen** | **Quality score** | **Red point score** | **Findnigs in MDD and BD patients compared to HC** |
| --- | --- | --- | --- | --- | --- | --- | --- | --- | --- | --- | --- | --- | --- | --- | --- |
|  |  |  |  |  | **Cases**  **(M/F)** | **Control**  **M/F** | **Total**  **M/F** | **Case-Mean (SD)** | | **Control- Mean**  **(SD)** |  |  |  |  |  |
| 1 | (Ahola et al. 2010) | Finland | Depression | ND | 214 (78/136) | 1012 (470/542) | 1226  (548/678) | 46.8 (12.3) | | 44.9(12.5) | TC,TG | Serum | 4.25 | 19 | TC#  TG# |
| 2 | (Aijanseppa et al. 2002) | Finland | Depressive symptom | ND | 64  (64/0) | 357  (357/0) | 421  (421/0) | 77.6 (5) | | 76 (4.6) | TC, HDL, LDL, TG | Serum | 4.5 | 21 | TC*  HDL*  LDL*  TG |
| 3 | (Akgün, Köken et al. 2017) | Turkey | BD with MetS | BD without Mets | 20  (5/15) | 20  (6/14) | 40  (11/29) | 46.4 (10.2) | | 39.8 (7.5) | HDL, TG | Serum | 7 | 10 | HDL*  TG# |
| 4 | (Xu, Zhao et al. 2024) | China | MDD | HC | 139 (64/75) | 65  (27/38) | 204  (91/113 | 37,2  (1,6) | | 36,3  (1,2) | TC | Serum | 4.5 | 18 | TC* |
| 5 | (Algul et al. 2013) | Turkey | MDD | HC | 20  (8/12) | 40  (18/22) | 60  (26/34) | 38.6 (3.3) | | 39.8 (2.0) | HDL,LDL,TG,TC | Serum | 5 | 19.5 | HDL  LDL  TG  TC |
| 6 | (Aliyazicioglu et al. 2011) | Turkey | MDD | HC | 78  (15/62) | 64  (24/35) | 142  (39/97) | 38 (11) | | 28 (9) | TC,TG,HDL,LDL, ApoA1, ApoB | Serum | 3 | 24 | TC#  TG  HDL  LDL#  Apo A  Apo B# |
| 7 | (Almeida-Montes et al. 2000) | Mexico | MDD | - | 33  (8/25) | - | 33  (8/25) | 31.95 (11.23) | | - | TC ,HDL,LDL  TG | Serum | 3 | 17.75 | TC ,HDL,LDL  TG |
| 8 | (Baek et al. 2014) | South Korea | MDD | - | 555  (150/405) | - | 555  (150/405) | 46.29 (15.54) | | - | TC, TG, HDL,LDL  VLDL | Serum | 4 | 19 | TC, TG, HDL,LDL,  VLDL |
| 9 | (Baghai, Varallo-Bedarida et al. 2010) | Germany | MDD | HC | 86 (53/33) | 80 (30/50) | 166  (83/83) | 49.9 (13.1) | | 50.6 (13.9) | TG, HDL | Serum | 4.5 | 14.5 | TG*  HDL# |
| 10 | (Baghai, Varallo-Bedarida et al. 2018) | Germany | MDD | HC | 100 (37/63) | 104 (47/57) | 204 (84/120) | 46.6 (14.8) | | 54.7 (14.4) | TC, LDL,TG  HDL | Blood | 5 | 9,5 | TC*  LDL  TG#  HDL* |
| 11 | (Bajwa et al. 1992) | USA | MDD | HC | 30 (17/13) | 30 (17/13) | 60  (34/26) | 38.2 (10.7) | | 36.2 (12.1) | TC | Plasma | 3.25 | 15.75 | TC# |
| 12 | (Bilici, Efe et al. 2001) | Turkey | MDD | HC | 18 (5/13) | 32 (16/16) | 50 (21/29) | 42.2 (9.7) | | 42.1 (7.4) | TG,TC | Plasma | 6 | 8,5 | TG#  TC# |
|  |  |  |  |  | 12 (4/8) |  |  | 40.4 (6.4) | |  |  |  |  |  |  |
| 13 | (Bortolasci, Vargas et al. 2014) | Brazil | BD | HC | 45 (12/33) | 199 (82/117) | 244 (94/150) | 44.5 (8.9) | | 46.4 (8.3) | HDL | Serum | 7 | 7,5 | HDL  PON1# |
|  |  |  |  |  |  |  |  |  |  |  | PON1 | Plasma |  |  |  |
|  |  |  | MDD |  | 91 (22/69) |  | 290 (104/186) | 47.0 (8.2) | |  | HDL | Serum |  |  | HDL  PON1# |
|  |  |  |  |  |  |  |  |  |  |  | PON1 | Plasma |  |  |  |
| 14 | (Ormonde do Carmo, Mendes-Ribeiro et al. 2015) | Brazil | MDD | HC | 22 (4/18) | 27 (10/17) | 49 (14/35) | 31 (2) | | 33 (2) | HDL,  LDL,  TG | Blood | 6 | 5 | HDL  LDL  TG# |
| 15 | (Cizza et al. 2009) | USA | MDD | HC | 77 (0/77) | 41 (0/41) | 118  (0/118) | 35.5 (7) | | 35.2 (7) | TG , TC, LDL,  HDL | Serum | 4.75 | 14.5 | TG#  TC#  LDL#  HDL* |
| 16 | (Das et al. 2010) | India | Unipolar depression | HC | 30 (12/18) | 30 (12/18) | 60 (24/36) | 41.1 (10.58) | | 42.03 (6.86) | TC, LDL,  HDL,TG, VLDL | Serum | 4.75 | 8.5 | TC  LDL  HDL  TG  VLDL |
| 17 | (van Reedt Dortland, Giltay et al. 2010) | Netherlands | MDD-current | HC | 761 (256/505) | 629 (243/386) | 1390  (499/891) | 41.7 (12.9) | | 41.20 (12.9) | HDL,TC,LDL,TG | Serum | 5.75 | 18.5 | HDL  TC  LDL  TG |
|  |  |  | MDD-remitted |  | 1071  (316/755) |  | 1700  (559/1141) | 42.4 (12.9) | |  |  |  |  |  |  |
| 18 | (Eidan et al. 2019) | Iraq | MDD | HC | 38  (24/14) | 30  (13/17) | 68  (37/31) | 30.76 (14.1) | | 31.1 (15.4) | TC,HDL,LDL,TG | Plasma | 4.75 | 16 | TC  HDL  LDL  TG# |
|  |  |  |  |  | 22  (16/6) |  | 52  (29/23) | 36.91 (10.3) | |  |  |  |  |  |  |
| 19 | (Ekinci et al. 2017) | Turkey | MDD | HC | 37  (15/22) | 50  (13/37) | 87  (28/59) | 42.17 (12.24) | | 43 (4.23) | LDL_1, TG_1, TC_1, HDL_1 | Serum | 5.25 | 9.5 | LDL_1  TG_1  TC_1  HDL_1 |
|  |  |  |  |  | 102  (27/75) |  | 152  (40/112) |  |  | 44.12 (4.23) | LDL_2, TG_2, TC_2, HDL_2 |  | 6.25 | 9.5 | LDL_1  TG_1  TC_1  HDL_1 |
|  |  |  |  |  | 139  (42/97) |  | 189  (55/134) |  |  | 44.12 (4.23) | LDL, TG, TC, HDL |  |  |  | LDL_1  TG_1  TC_1  HDL_1 |
| 20 | (Enko, Brandmayr et al. 2018) | Austria | MDD | HC | 94  (30/64) | 152  (58/94) | 246  (88/158) | - | | - | HDL,TG,TC,  LDL | Plasma | 5 | 12 | HDL*  TG#  TC  LDL |
| 21 | (Ergun et al. 2004) | Turkey | Major depression, Dysthymia, Not specified depression | ND | 42  (14/28) | 147  (68/79) | 189  (82/107) |  | |  | TC, HDL, LDL,TG | Plasma | 4.75 | 17.5 | TC  HDL  LDL  TG |
| 22 | (Guidara, Messedi et al. 2021) | Tunisia | BD | HC | 33 (33/0) | 40 (40/0) | 73 (73/0) | 33.69 (10.26) | | 35.96  (7.30) | TC, TG,  HDL, LDL | Plasma | 6 | 7 | TC  TG  HDL  LDL |
| 23 | (Hamidifard et al. 2008) | Iran | MDD | HC | 35 (6/29) | 35 (6/29) | 70 (12/58) | 35.2 (10.72) | | 33.71  (10.21) | TC, TG,  HDL, LDL, Lipoprotein_A | Plasma | 5.25 | 16.25 | TC*  TG  HDL  LDL*  Lipoprotein_A# |
| 24 | (Heckbert et al. 2010) | USA | Major depression | ND | 448 (185/263) | 2995 (1611/1384) | 3448 (1796/1647) | 60 (13.1) | | 64.8 (12.3) | LDL | Blood | 2.75 | 20.5 | LDL |
| 25 | (Hocaoglu, Kural et al. 2012) | Turkey | MDD | HC | 30 (6/24) | 30 (16/14) | 60 (22/38) | 38 (13) | | 30 (9) | TC,TG,HDL,  LDL | Serum | 2.5 | 21 | TC#  TG  HDL#  LDL |
| 26 | (Huang and Chen 2004) | Taiwan | MDD | HC | 68 (31/37) | 39 (20/19) | 107 (51/56) | 43.5 (14.0) | | 50.2 (11.2) | TC,TG,HDL,  VLDL,LDL, CAST1, CAST2 | Serum | 3.25 | 21.5 | TC  TG  HDL  VLDL  LDL  CAST 1  CAST 2 |
| 27 | (Huang 2005) | Taiwan | MDD | HC | 109 (32/77) | 59 (22/37) | 168 (54/114) | 31.4 (8.5) | | 29.5 (4.4) | TC,TG,HDL,  LDL, CAST1, CAST2, VLDL | Serum | 3.75 | 23.5 | TC  TG*  HDL#  LDL  CAST 1  CAST 2, VLDL* |
| 28 | (Hui, Yin et al. 2019) | China | BD | HC | 37 (15/22) | 37 (15/22) | 74 (30/44) | 29.78 (10.05) | | 29.95 (9.02) | HDL | Serum | 5 | 8 | HDL * |
| 29 | (Jirakran, Vasupanrajit et al. 2023) | Thailand | MDD | HC | 66  (18/48) | 67  (9/58) | 133  (27/106) | 36.9 (11.5) | | 37.9 (9.2) | LCAT activity,  HDL, Apo A, Apo B, TC,TG,LDL, Castelli1, TG/HDL | Serum | 7 | 9.5 | LCAT activity  HDL  Apo A  Apo B  TC  TG  LDL  Castelli1  TG/HDL |
| 30 | (Jow et al. 2006) | Taiwan | MDD | HC | 69  (34/35) | 51  (25/26) | 120  (59/61) | 28.7 (7.8) | | 27.8 (7.6) | TC | Serum | 4.5 | 15 | TC |
| 31 | (Kahl, Schweiger et al. 2015) | Germany | MDD-current | HC | 10 (3/7) | 90 (57/33) | 100 (60/40) | 51.3 (14.7) | | 52.3 (14.9) | HDL,TG | Plasma | 5 | 9.5 | HDL  TG |
| 32 | (Kale et al. 2014) | India | Depressive symptom | HC | 40 (15/25) | 30 (14/16) | 70 (29/41) | 35.44 (1.23) | | 32.43 (2.13) | TG,HDL,LDL,VLDL,TC | Serum | 2 | 22.5 | TG*  HDL*  LDL*  VLDL*  TC* |
| 33 | (Karadeniz, Yaman et al. 2020) | Turkey | MDD | HC | 37 (11/26) | 33 (12/21) | 70 (23/47) | 14.62 (1.60) | | 14.72 (1.15) | HDL,TC,TG | Serum | 6 | 7 | HDL  TC*  TG |
| 34 | (Karlović, Buljan et al. 2004) | Croatia | MDD (only) | HC | 38 (38/0) | 39 (39/0) | 77 (77/0) | 46.2 (11.3) | | 43.8 (10.1) | TC,TG,LDL,  HDL,CAST2 | Serum | 5 | 17.5 | TC*  TG*  LDL#  HDL*  CAST 2* |
|  |  |  | MDD (+PTSD) |  | 37 (37/0) |  | 76 (76/0) | 42.9 (7.3) | |  |  |  |  |  |  |
| 35 | (Kasak, Ceylan et al. 2022) | Turkey | BD | HC | 39 (12/27) | 36 (9/27) | 75 (21/54) | 16.7 (1.27) | | 17.0 (0.75) | HDL,TG | Serum | 6 | 7.5 | HDL  TG# |
| 36 | (Kennedy, Islam et al. 2021) | Canada | BD | HC | 55 (20/35) | 47 (23/24) | 102 (43/59) | 17.64 (1.76) | | 17.34 (1.69) | HDL,TC,LDL,TG | Blood | 4 | 10.5 | HDL*  TC#  LDL#  TG# |
| 37 | (Khalid et al. 1998) | India | MDD | HC | 28 (15/13) | 28 (15/13) | 56  (30/26) | 38.64 (10.74) | | 39.25 (10.88) | TC, TG, HDL, LDL | Serum | 3.75 | 14.5 | TC  TG*  HDL#  LDL* |
| 38 | (Kim et al. 2004) | South Korea | Depressive symptom | HC | 149 (62/87) | 251 (101/150) | 400  (163/237) | 40.5 (18.1) | | 38 (15.9) | TC_1, TC_2 | Serum | 4.25 | 18.5 | TC_1*  TC_2* |
|  |  |  |  |  |  |  |  | 42 (16.5) | |  |  |  |  |  |  |
| 39 | (Kodydková, Vávrová et al. 2009) | Czech Republic | Depressive disorders | HC | 35 (0/35) | 35 (0/35) | 70 (0/70) | 63.11 (19.40) | | 65.07 (18.39) | TC,TG,HDL,LDL,APOA,APOB, | Serum | 3 | 19 | TC  TG#  HDL  LDL  APO A  APO B  PON1 |
|  |  |  |  |  |  |  |  |  |  |  | PON1 |  | 3.5 | 20.5 |  |
| 40 | (Koponen et al. 2015) | Finland | MDD + Dysthymia | - | 448 (136/312) | 0 | 448 (136/312) | 51 (10) | | - | TC,HDL,LDL,TG | Serum | 3.5 | 23.5 | TC  HDL  LDL  TG |
| 41 | (Kotan, Sarandol et al. 2011) | Turkey | MDD | HC | 50 (11/39) | 44 (10/34) | 94 (21/73) | 33.1 (10.0) | | 33.2 (7.9) | TC,LDL,VLDL,HDL,CAST1,CAST2,TG,APOA,APOB, OxLDL,VitE, PON1 | Serum | 3.75 | 14 | TC,LDL,VLDL,HDL,CAST1,CAST2,TG,APOA,APOB, OxLDL,VitE, PON 1 |
| 42 | (Lehto, Niskanen et al. 2010) | Finland | Depressive symptom | ND | 269 (269/0) | 2187 (2187/0) | 2456 (2456/0) | 53.49 (4.59) | | 53.01(5.1) | TC,LDL,HDL,TG,ApoB, LDL/ApoB | Serum | 6.25 | 16 | TC  LDL#  HDL*  TG#  Apo B  LDL/ApoB |
|  |  |  | MDD (≥3yr.duration) | HC | 43 (19/24) | 88 (39/49) | 131 (58/73) | 50.56 (4.8) | | 49.86(8.01) | TC_1,HDL_1, LDL_1,TG_1, CAST1_1, CAST2_1, Non-HDL_1 |  | 6.5 | 17 | TC_1  HDL_1*  LDL_1#  TG_1#  CAST1_1*  CAST2_1*  Non-HDL_1* |
|  |  |  |  |  | 45 (20/25) |  | 133 (59/74) | 49.09 (6.63) | |  | TC_2,HDL_2, LDL_2,TG_2, CAST1_2, CAST2_2, Non-HDL_2 |  |  |  | TC_2  HDL_2*  LDL_2#  TG_2#  CAST1_2*  CAST2_2*  Non-HDL_2* |
| 43 | (Lehto, Hintikka et al. 2008) | Finland | Depressive symptom | ND | 63 (19/44) | 61 (17/44) | 124 (36/88) | 54.68 (8.59) | | 55.59 (8.96) | TC,HDL,LDL,TG,CAST2,CAST1 | Serum | 5.75 | 17.5 | TC  HDL*  LDL  TG  CAST 2#  CAST 1# |
| 44 | (Liu et al. 2015) | China | MDD | HC | 135 (70/65) | 111 (56/55) | 246(126/120) | 38.87 (11.87) | | 39.15 (11.74) | TC,TG, LDL, ApoA,ApoB,  HDL | Plasma | 3.75 | 20.5 | TC  TG#  LDL  Apo A  Apo B  HDL |
| 45 | (Maes, Delanghe et al. 1994) | Belgium | Depressed mood in Dysthymic disorder + Adjustment disorder | HC | 16 (16/0) | 26 (26/0) | 42 (42/0) | 45.0 (10.0) | | 40.0 (14.9) | LCAT activity,TC | Serum | 7 | 9.5 | LCAT activity,TC |
|  |  |  | MDD (witout melancholia) |  | 14 (14/0) |  | 40 (40/0) | 51 (9.7) | |  |  |  |  |  |  |
|  |  |  | MDD (with melancholia) |  | 17 (17/0) |  | 43 (43/0) | 53.1 (12.3) | |  |  |  |  |  |  |
| 46 | (Maes, Smith et al. 1997) | Belgium | MDD | HC | 36 (5/31) | 28 (5/23) | 64 (10/54) | 51.1 (13.7) | | 47.7 (14.2) | TC,TG,HDL,LDL,VitE | Serum | 6.75 | 9.5 | TC*  TG#  HDL*  LDL  VitE |
| 47 | (Vaghef-Mehrabani, Izadi et al. 2021) | Iran | MDD | ND | 75 (0/75) | 150 (0/150) | 225 (0/225) | 39.64 (7.70) | | 39.97 (7.58) | TG,TC,LDL,HDL | Serum | 6 | 16 | TG#  TC#  LDL#  HDL* |
| 48 | (Segoviano-Mendoza, Cárdenas-de la Cruz et al. 2018) | Mexico | MDD | HC | 204 (35/169) | 206 (40/166) | 410 (75/335) | 37.3 (10.0) | | 36.8 (6.6) | HDL,TC,LDL,VLDL,TG | Serum | 5 | 18 | HDL  TC*  LDL*  VLDL#  TG# |
|  |  |  |  |  | 202 (38/171) |  | 467 (76/337) |  |  |  |  |  | 6.5 | 20 |  |
|  |  |  | MDD-suicidal |  | 59 (17/42) |  | 265 (57/208) | 35.2 (10.5) | |  |  |  | 5 | 18 |  |
| 49 | (Messaoud, Mensi et al. 2017) | Tunisia | MDD | HC | 162 (54/108) | 151 (50/101) | 313 (104/209) | 39.67 (9.98) | | 38.92 (13.28) | TC | Plasma | 4.25 | 18.5 | TC |
|  |  |  |  |  | 110 (35/75) |  | 261 (85/176) | 44.33 (10.50) | |  | HDL,TC,TG,LDL  HDL |  |  |  | HDL  TC  TG  LDL  HDL |
|  |  |  | MDD-suicidal |  | 52 (19/33) |  | 203 (69/134) | 29.84 (8.78) | |  |  |  |  |  |  |
| 50 | (Moreira, Jansen et al. 2017) | Brazil | MDE in MDD | ND | 77 (13/64) | 863 (383/480) | 940 (396/544) | 26.09 (2.22) | | 25.81 (2.17) | TG,TC,HDL,LDL,CAST1,CAST2 | Serum | 6 | 8 | TG  TC  HDL*  LDL  CAST 1#  CAST 2 |
|  |  |  | MDE in BD |  | 32 (6/26) |  | 895 (389/506) | 25.59 (2.24) | |  |  |  |  |  |  |
| 51 | (Nakao et al. 2004) | Japan | MDE | ND | 24 (24/0) | 963 (963/0) | 987 (987/0) | 36 (10) | | 35 (10) | TG,TC,HDL | Serum | 4.5 | 20.5 | TG  TC#  HDL |
| 52 | (Nunes et al. 2015) | Brazil | Mood disorders (i.e., MDD and BD) | HC | 62 (51/11) | 113 (69/44) | 175 (120/55) | 46.5 (6.9) | | 45.2 (8.1) | PON1 | Plasma | 6 | 9.5 | PON1* |
|  |  |  |  |  | 134 (34/100) | 197 (82/115) | 331 (116/215) | 46.2 (8.6) | | 46.4 (8.3) | TC,HDL,LDL,TG | Serum | 7 | 7 | TC  HDL  LDL  TG |
| 53 | (Olie et al. 2011) | France | MDD + Adjustment disorder | ND | 785 (234/551) | 2422 (1291/1131) | 3207(1525/1682) | 39 (15.16) | | 54.28 (15.83) | TC,TG | Serum | 3.5 | 21 | TC  TG |
| 54 | (Olusi and Fido 1996) | Kuwait | MDD | HC | 100 (64/36) | 100 (64/36) | 200 (128/72) | 39.58 (10.2) | | 39.96 (9.8) | TC,TG,HDL,LDL,APOA1,APOB | Serum | 4.25 | 16.5 | TC*  TG  HDL#  LDL*  APO A  APO B |
| 55 | (Onuegbu et al. 2007) | Nigeria | MDD | HC | 60 (22/38) | 40 (19/21) | 100 (41/59) | 40.3 (12.3) | | 40.1 (10.1) | TC,TG,HDL,LDL | Plasma | 4 | 18.5 | TC  TG#  HDL  LDL |
| 56 | (Oxenkrung et al. 1983) | USA | MDD in female (18-59 yr.) | HC | 24(0/24) | 24(0/24) | 48(0/48) | 39.8 (12.73) | | 40.2 (18.61) | TC_1, TC_2, TC_3, TC_4 | Serum | 2.5 | 23.5 | TC_1, TC_2, TC_3, TC_4 |
|  |  |  | MDD in male (18-59 yr.) |  | 24(24/0) | 24(24/0) | 48(48/0) | 39.9 (11.75) | | 42.3 (17.63) |  |  |  |  |  |
|  |  |  | MDD in female (60-85 yr.) |  | 24(0/24) | 24(0/24) | 48(0/48) | 67.8 (3.91) | | 67.2 (3.91) |  |  |  |  |  |
|  |  |  | MDD in male (60-85 yr.) |  | 24(24/0) | 24(24/0) | 48(48/0) | 67 (5.87) | | 69.4 (4.40) |  |  |  |  |  |
| 57 | (Palta et al. 2014) | USA | Depressive symptom | ND | 218 (42/176) | 346 (131/215) | 564 (173/391) | 72.5 (6.3) | | 73 (6.6) | TC,TG,HDL | Blood | 3.25 | 20 | TC  TG  HDL |
| 58 | (Park et al. 2013) | South Korea | MDD | - | 201 (86/115) | - | 201 (86/115) | 53.6 (15.2) | | - | TC,TG,HDL | Serum | 2 | 25 | TC,TG,HDL |
| 59 | (Partonen et al. 1999) | Finland | Depressive symptom | ND | 4314(4314/0) | 24790 (24790/0) | 29104 (29104/0) | - | | - | TC, HDL | Serum | 4.25 | 20 | TC*  HDL# |
| 60 | (Patra, Khandelwal et al. 2014) | India | MDE | HC | 30 (19/11) | 30 (19/11) | 60 (38/22) | 32.47 (11.41) | | 32.63 (11.20) | TC,LDL,HDL,TG | Serum | 3.5 | 20.5 | TC*  LDL*  HDL  TG |
| 61 | (Peng et al. 2016) | China | MDD | HC | 228 (43/185) | 251(50/201) | 479(93/386) | 39.55 (8.06) | | 39.97 (7.37) | HDL,LDL,TC,TG | Blood | 4 | 11 | HDL#  LDL  TC  TG |
| 62 | (Peng et al. 2017) | China | MDD | HC | 305 (66/239) | 312 (172/140) | 617 (238/379) | 39.9(8.47) | | 33.8 (9.86) | TC,LDL,HDL,TG | Serum | 6 | 12 | TC#  LDL#  HDL  TG |
| 63 | (Peterfalvi_1 et al. 2019) | Hungary | MDD | HC | 42 (9/33) | 20 (7/13) | 62 (16/46) | 35.4(9.73) | | 35.8(8.53) | TC,TG,HDL,LDL,CAST2,CAST1 | Serum | 4.25 | 13 | TC  TG  HDL  LDL  CAST 2  CAST 1 |
| 64 | (Rahiminejad et al. 2014) | Iran | Depressive symptom | ND | 38 (0/38) | 82 (0/82) | 120 (0/120) | 24.8(4.6) | | 23.6(4.7) | TC,LDL,HDL,TC |  | 2.75 | 18.5 | TC  LDL  HDL  TC |
| 65 | (Ranjekar, Hinge et al. 2003) | India | BD | HC | 10 (10/0) | 31 (31/0) | 41 (41/0) | 40.8 (8.29) | | 40.9 (8.84) | HDL,ApoA1,ApoB,TC,LDL,VLDL,TG | Plasma | 6 | 8 | HDL*  Apo A  Apo B  TC  LDL  VLDL#  TG# |
| 66 | (Qi, Wang et al. 2024) |  |  |  |  |  |  |  | |  |  |  |  |  |  |
| 67 | (Hamidreza Roohafza et al. 2005) | Iran | MDD | HC | 25 (5/20) | 25 (7/18) | 50 (12/38) | 33.2 (6.1) | | 33.2 (7.7) | HDL,TC,TG,LDL | Serum | 3.5 | 13.5 | HDL*  TC* TG* LDL* |
| 68 | (Ruljancic et al. 2011) | Croatia | MDE | HC | 132 (56/76) | 97 (48/49) | 229 (104/125) | 44 (22-57)  44 (20-59) | | 36 (21-59) | TC | Serum | 3.25 | 20 | TC* |
| 69 | (Rybka, Kędziora-Kornatowska et al. 2013) | Poland | Depressive disorders | HC | 15 (15/0) | 19 (19/0) | 34 (34/0) | 59.7 (1.91) | | 62.3 (2.84) | TC,TG | Serum | 6 | 6.5 | TC#  TG# |
| 70 | (Sadeghi, Roohafza et al. 2011) | Iran | MDD | HC | 153 (90/63) | 147 (69/78) | 300 (159/141) | 31.21 (10.41) | | 32.00 (8.21) | TC,LDL,HDL,ApoA,ApoB | Serum | 4.25 | 17.5 | TC#  LDL#  HDL*  Apo A*  Apo B# |
| 71 | (Sagud, Mihaljevic-Peles et al. 2009) | Croatia | MDD | HC | 34 (0/34) | 50 (0/50) | 84 (0/84) | 50.1 (6.6) | | 44.7 (12.8) | CAST1,CAST2,LDL,TC | Serum | 4.75 | 17.5 | CAST 1#  CAST 2#  LDL#  TC# |
|  |  |  | BD (depressive episode) |  | 19 (0/19) |  | 69 (0/69) | 44.1 (12.9) | |  | CAST1_1,CAST2_1,LDL_1,TC_1 |  |  |  | CAST 1#  CAST 2#  LDL#  TC# |
| 72 | (Sanyal et al. 2000) | India | MDD | HC | 13 (8/5) | 13 (8/5) | 26 (16/10) | - | | - | TC | Serum | 2.75 | 15 | TC |
| 73 | (Sarandol, Sarandol et al. 2006) | Turkey | MDD | HC | 86 (24/62) | 36 (10/26) | 122 (34/88) | 40.5 (10.5) | | 37.2 (7.3) | TC,HDL,LDL,VLDL,CAST1,TG,ApoB,ApoA1 | Serum | 3.25 | 15 | TC#  HDL#  LDL#  VLDL  CAST 1  TG  Apo B#  Apo A* |
| 74 | (Scharnholz et al. 2014) | Germany | MDD | HC | 20 (7/13) | 34 (19/15) | 54 (26/28) | 40.3 (12.5) | | 45.1 (14.5) | TC | Serum | 3.75 | 20 | TC |
|  |  |  |  |  |  |  |  |  |  |  | HDL |  | 4.25 | 25 | HDL* |
|  |  |  |  |  |  |  |  |  |  |  | TG |  | 4.75 | 30 | TG |
| 75 | (Sevincok, Buyukozturk et al. 2001) | Turkey | MDD | HC | 27 (7/20) | 24 (6/18) | 51 (13/38) | 33.29 (6.12) | | 33.20 (7.78) | TC,TG,HDL,LDL | Serum | 6 | 7 | TC  TG#  HDL*  LDL# |
| 76 | (Shao et al. 2017) | China | MDD | HC | 115 (45/70) | 115 (52/67) | 234 (97/137) | 37.76 (12.48) | | 37.45 (12.35) | TG | Serum | 5.5 | 18.25 | TG# |
|  |  |  |  |  | 97 | 113 | 210 |  |  |  |  |  |  |  |  |
| 77 | (Shapiro, Kennedy et al. 2022) | Canada | BD | HC | 88 (31/57) | 89 (39/50) | 177 (70/10) | 17.5 (1.7) | | 17.2 (1.8) | HDL,TC,LDL,TG | Blood | 6 | 7 | HDL  TC#  LDL#  TG# |
| 78 | (Su, Li et al. 2019) | China | Unipolar depression | HC | 195 (49/146) | 89 (25/64) | 284 (74/210) | 44.10 (13.94) | | 47.34 (13.05) | TG,TC,HDL,LDL,VLDL | Serum | 3.75 | 20.5 | TG*  TC*  HDL  LDL*  VLDL* |
|  |  |  | Bipolar depression |  | 92 (42/50) |  | 181 (67/114) | 24.76 (12.75) | |  |  |  |  |  |  |
| 79 | (Suaad et al. 2019) | Iraq | MDD | HC | 60 (40/20) | 30 (13/17) | 90 (53/37) | 30.76 (14.1) | | 31.1 (15.4) | TC,HDL,LDL,TG | Serum | 4 | 23 | TC  HDL  LDL  TG# |
| 80 | (Sonal Sukreet and Chaturvedi 2011) | India | BD | HC | 100 (58/42) | 52 (26/26) | 152 (84/68) | 63 (12) | | 52 (12) | TC,HDL,TG,LDL | Serum | 4 | 22 | TC#  HDL  TG#  LDL# |
| 81 | (Sullivan et al. 1994) | New Zealand | MDD | - | 90 (39/51) | - | 90 (39/51) | 31.31 (9.77) | | - | TC,TG | Blood | 0.5 | 24.5 | TC  TG |
| 82 | (Tuncel et al. 2016) | Turkey | MDD | HC | 30 (7/23) | 30 (7/23) | 60 (14/46) | 14.7 (2.03) | | 14.7 (2.03) | TG,TC | Serum | 7 | 8.5 | TG  TC# |
| 83 | (Tuncel et al. 2017) | Turkey | BD (Manic episode) | HC | 30 (11/19) | 30 (11/19) | 60 (22/38) | 34.4 (10.3) | | 35 (11.3) | HDL,TG,LDL | Serum | 6 | 9 | HDL*  TG  LDL* |
| 84 | (Vargas, Nunes et al. 2014) | Brazil | MDD | HC | 92 (22/70) | 201 (82/119) | 293 (104/189) | 47 (8.20) | | 46.37 (8.26) | TC,HDL,LDL,CAST1,CAST2,TG | Serum | 6 | 7 | TC  HDL*  LDL  CAST 1#  CAST 2#  TG |
|  |  |  | BD (depressive episode) |  | 49 (12/37) |  | 250 (94/156) | 44.53 (8.99) | |  |  |  |  |  |  |
| 85 | (Wagner, Musenbichler et al. 2019) | Germany | MDE | HC | 130 (61/69) | 61 (30/31) | 191 (91/100) | 44.24 (15.74) | | 43.05 (17.46) | HDL,TG.TC.LDL.LDL/HDL | Serum | 7 | 7 | HDL  TG#  TC  LDL#  LDL/HDL# |
| 86 | (Wei, Wang et al. 2022) | China | BD (depressive episode) | HC | 1994(1002/942) | 5810 (2850/2960) | 7754 (3852/3902) | 39.05 (0.30) | | 38.54 (0.145) | HDL,TC,TG,LDL,Apo B | Blood | 6 | 9.5 | HDL*  TC  TG#  LDL  Apo B |
| 87 | (Zhang et al. 2020) | China | MDD | HC | 49 (18/31) | 50 (20/30) | 99 (38/61) | 42.29 (10.55) | | 42.58 (12.27) | TC,TG,HDL | Serum | 7.5 | 15.5 | TC#  TG#  HDL* |
| 88 | (Draghici 2022) | Romania | Mood disorder | HC | 134 (19/115) | 108 (14/94) | 242 (33/209) | 65.67 (6.56) | | 65.25 (7.17) | TC,TG,HDL,LDL | Serum | 6 | 9 | TC  TG#  HDL*  LDL |
| 89 | (Chen, Sun et al. 2024) | China | MDD | HC | 138  (53/85) | 50  (19/31) | 188  (72/116) | 37.4(12.7) | | 36.5(12.2) | HDL,LDL,TC,TG | Serum | 7 | 10 | HDL#,LDL*,TC*,TG* |
| 90 | (Maes, Zhou et al. 2024) | Thailand | MDD | HC | 66  (18/48) | 67  (9/58) | 133  (27/106) | 37(12) | | 37(9) | Apo B/Apo A, ApoA, ApoB,CER(, Free CholesteroHDL, LDL,TC, TG,TG/HDL | Serum | 8 | 9 | Apo B/Apo A*, ApoA#, ApoB*,CER#, Free Cholesterol*, HDL#, LD*L,TC*, TG*,TG/HDL* |
| 91 | (Khalfan, Campisi et al. 2023) | Canada | MDD | HC | 186  (48/138) | 57  (29/28) | 234  (77/166) | 15(2) | | 15,1(2) | HDL#,LDL#,LDL/HDL,TC,TG,TG/HDL | Plasma | 5 | 17 | HDL,LDL,LDL/HDL#,TC*,TG*,TG/HDL* |

*: Indicates that patients have reduced levels of the measured metabolites compared to healthy control

#: Indicates that patients have increased levels of the measured metabolites compared to healthy control

MDD: Major depressive disorder, HC: Healthy controls,HDL: High-density lipoprotein, LDL: Low-density lipoprotein, TG: Triglyceride, TC: Total cholesterol, VLDL: very low-density lipoprotein, Apo: Apolipoprotein, CAST: castelli index, LCAT: Lecithin cholesterol acyltransferase

**ESF, Table 6. Results of meta-regression**

| **Variables** | **No. of Studies** | **Covariates** | **1-sided p-value** | **Z-Value** |
| --- | --- | --- | --- | --- |
| Castelli Risk Index 1 | 18 | Absence of MetS | 0.051 | -1.55 |
|  | 76 | Red Points | 0.009 | -2.35 |
| Castelli risk index 2 | 68 | Fasting | 0.021 | 2.02 |
|  | 17 | Caucasian% | 0.049 | 1.60 |
|  | 18 | Absence of MetS | 0.010 | -2.31 |
| Atherogenic index of plasma | 65 | Female | 0.022 | -2 |
| ApoB/ApoA | 12 | Acute & Remission | 0.000 | 4.02 |
| (TG+LDL+VLDL)/HDL+ApoA | 66 | Red Points | 0.018 | -2.08 |
| Total cholesterol | 72 | Acute & Remission | 0.040 | 1.74 |
| LDL | 14 | Absence of MetS | 0.023 | -1.99 |
|  | 56 | Acute & Remission | 0.012 | 2.24 |
|  | 48 | Age | 0.043 | 1.71 |
|  | 60 | Latitude | 0.024 | 1.96 |
|  | 58 | Red points | 0.036 | -1.80 |
| HDL | 17 | Absence of MetS | 0.035 | 1.80 |

Very low-density lipoprotein (VLDL), low-density lipoprotein (LDL), apolipoprotein A (ApoA), Castelli risk index-I = TC/HDL, Castelli risk index-II = LDL/HDL.

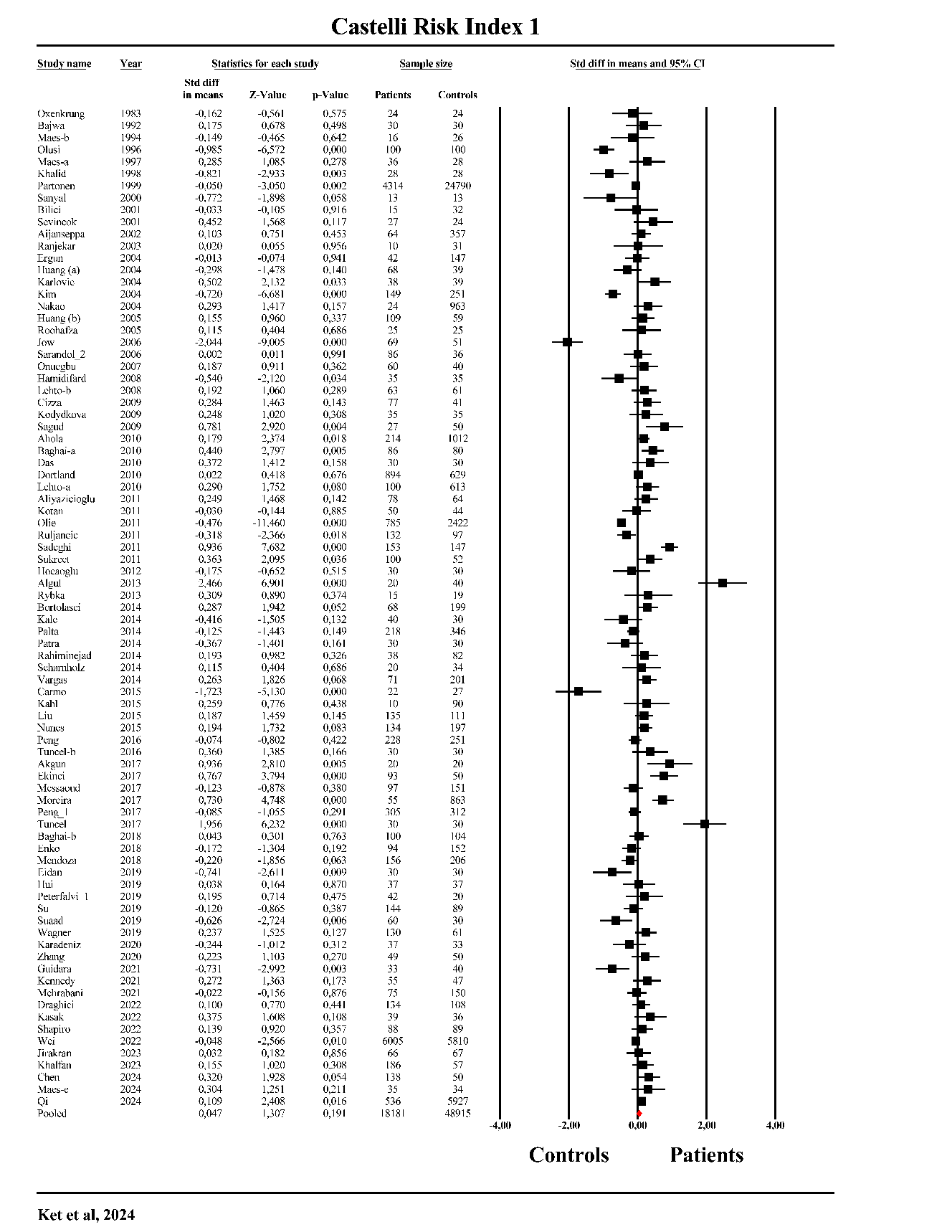

**ESF, Figure 1.** The forest plot of Castelli Risk Index 1 in patients with major depression as compared to healthy controls.

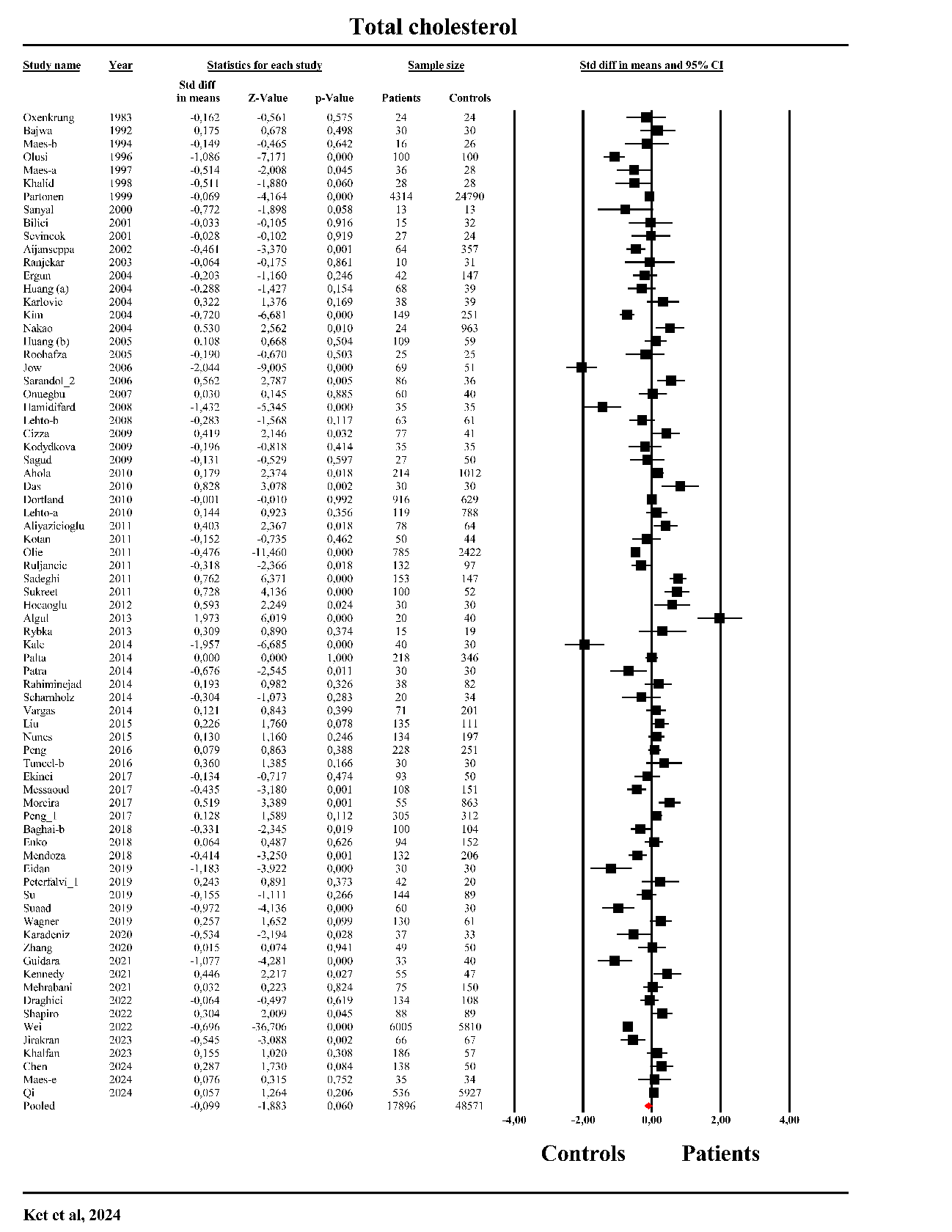

**ESF, Figure 2.** The forest plot of total cholesterol in patients with major depression as compared to healthy controls.

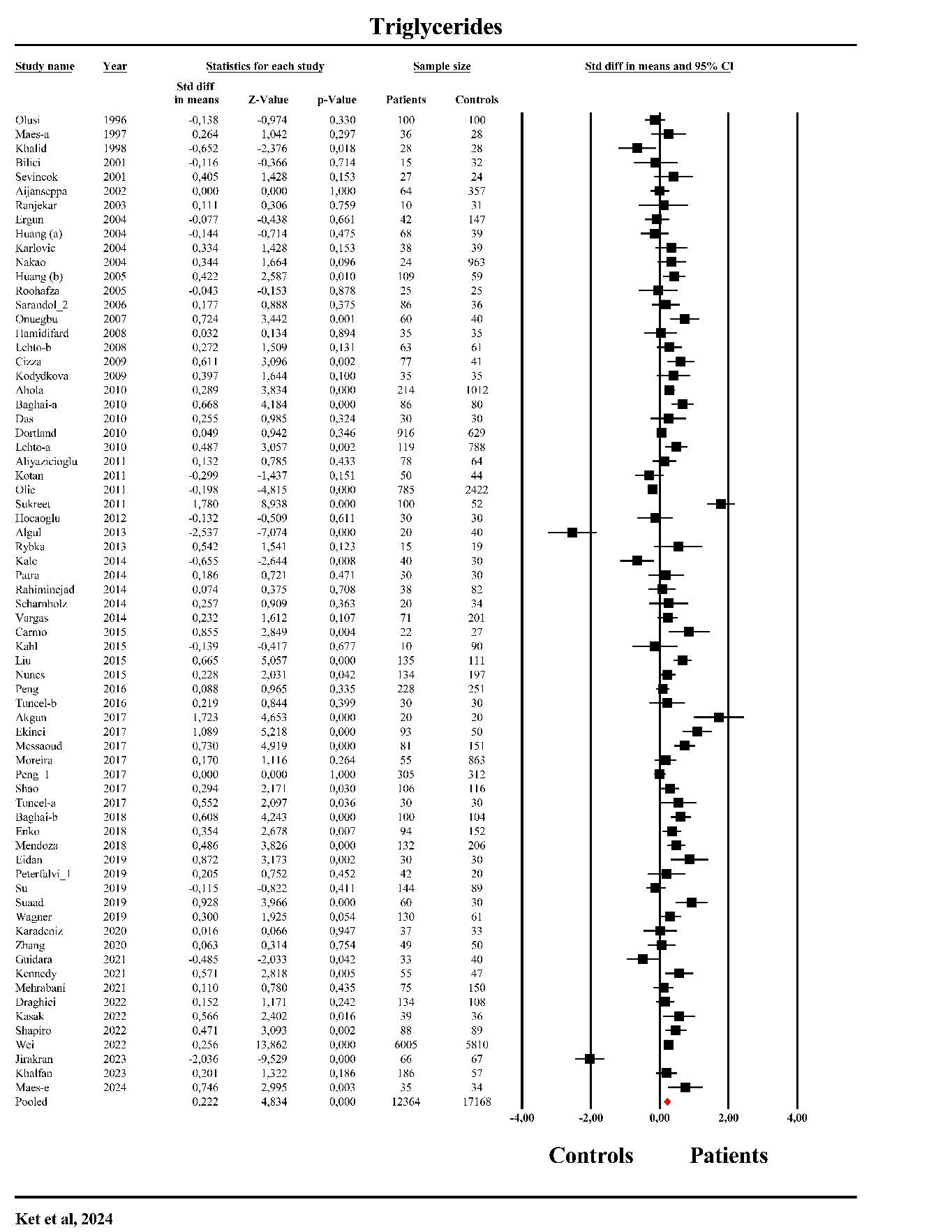

**ESF, Figure 3.** The forest plot of triglycerides in patients with major depression as compared to healthy controls.

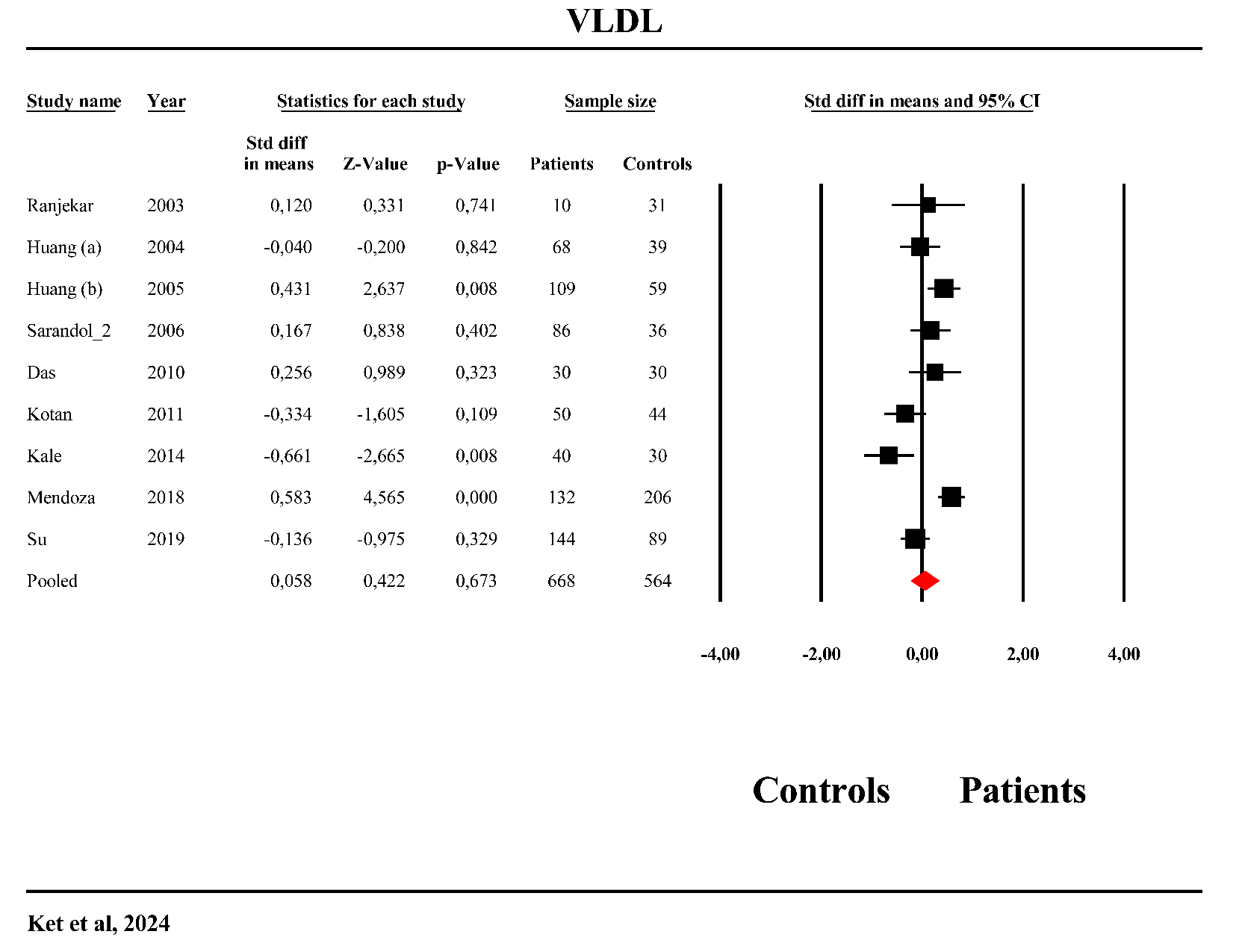

**ESF, Figure 4.** The forest plot of very low-density lipoprotein in patients with major depression as compared to healthy controls.

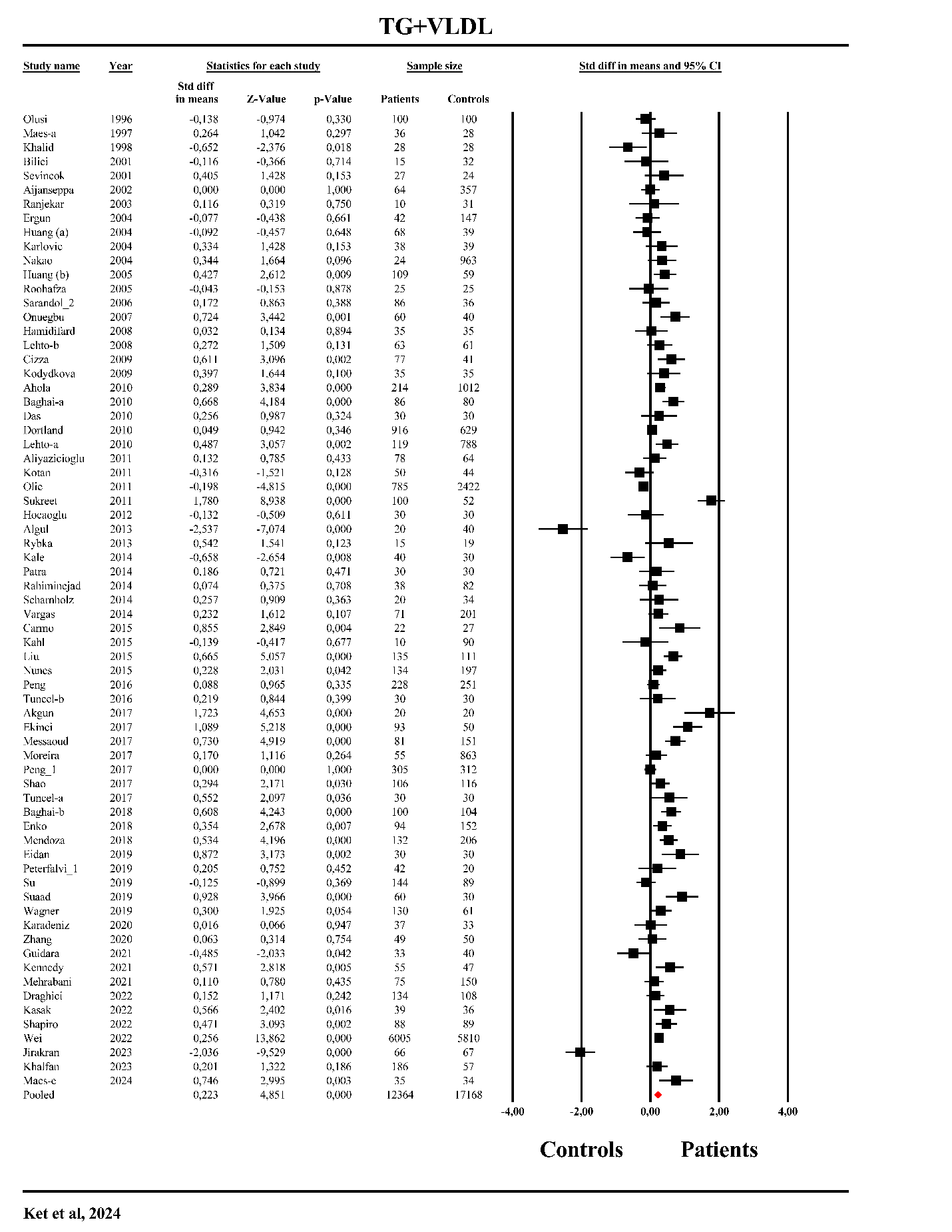

**ESF, Figure 5.** The forest plot of triglycerides (TG)+ very low-density lipoprotein (VLDL) in patients with major depression as compared to healthy controls.

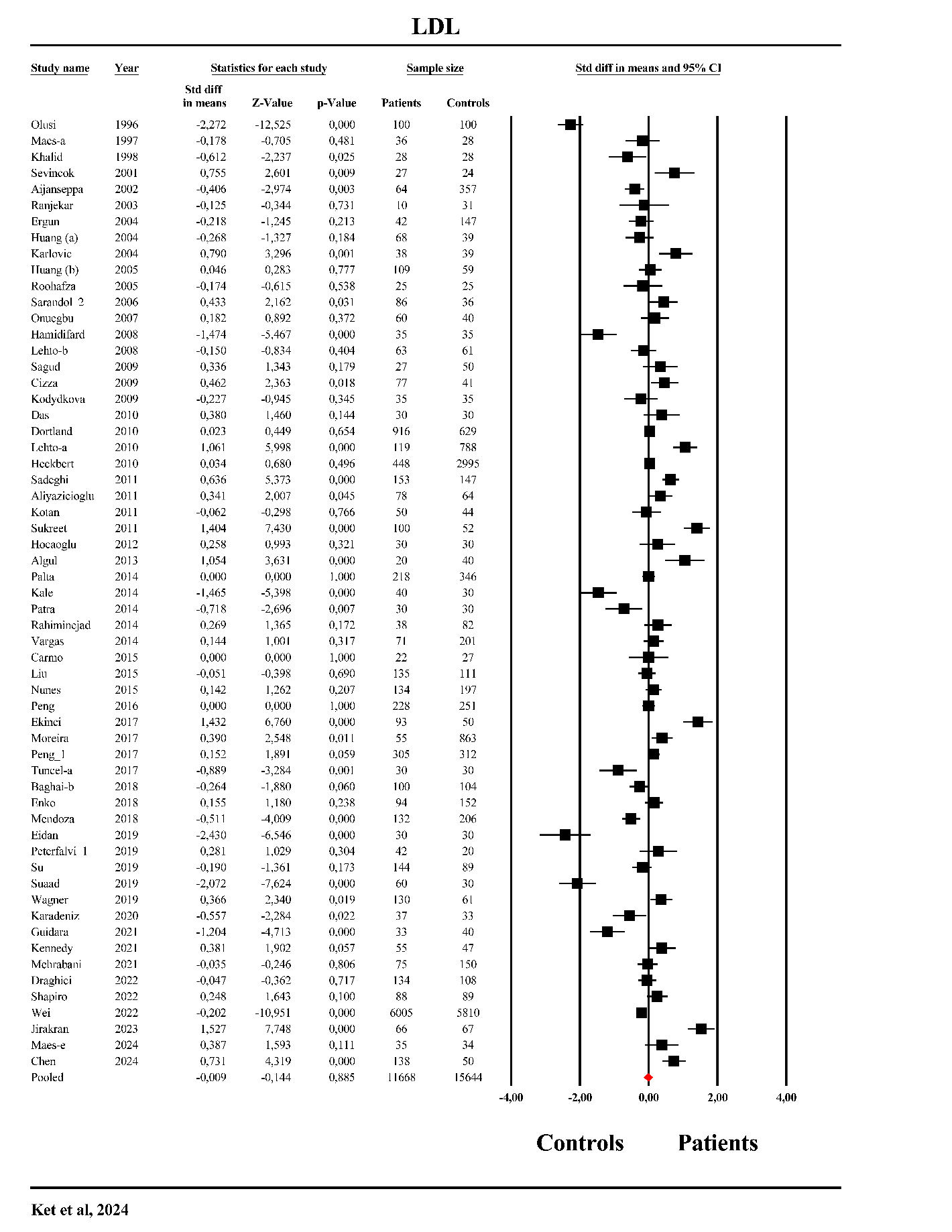

**ESF, Figure 6.** The forest plot of low-density lipoprotein (LDL) in patients with major depression as compared to healthy controls.

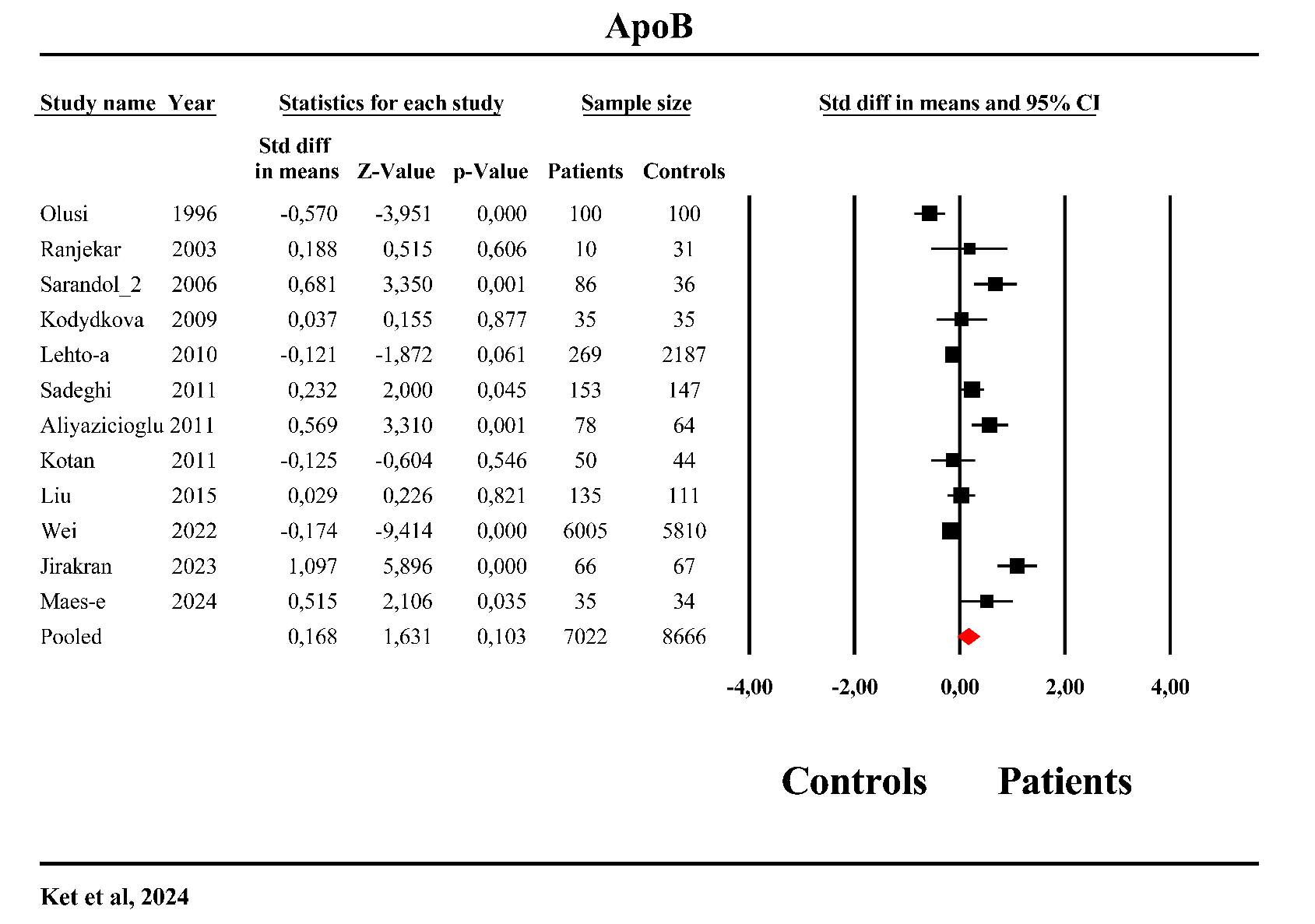

**ESF, Figure 7.** The forest plot of apolipoprotein (Apo)B in patients with major depression as compared to healthy controls.

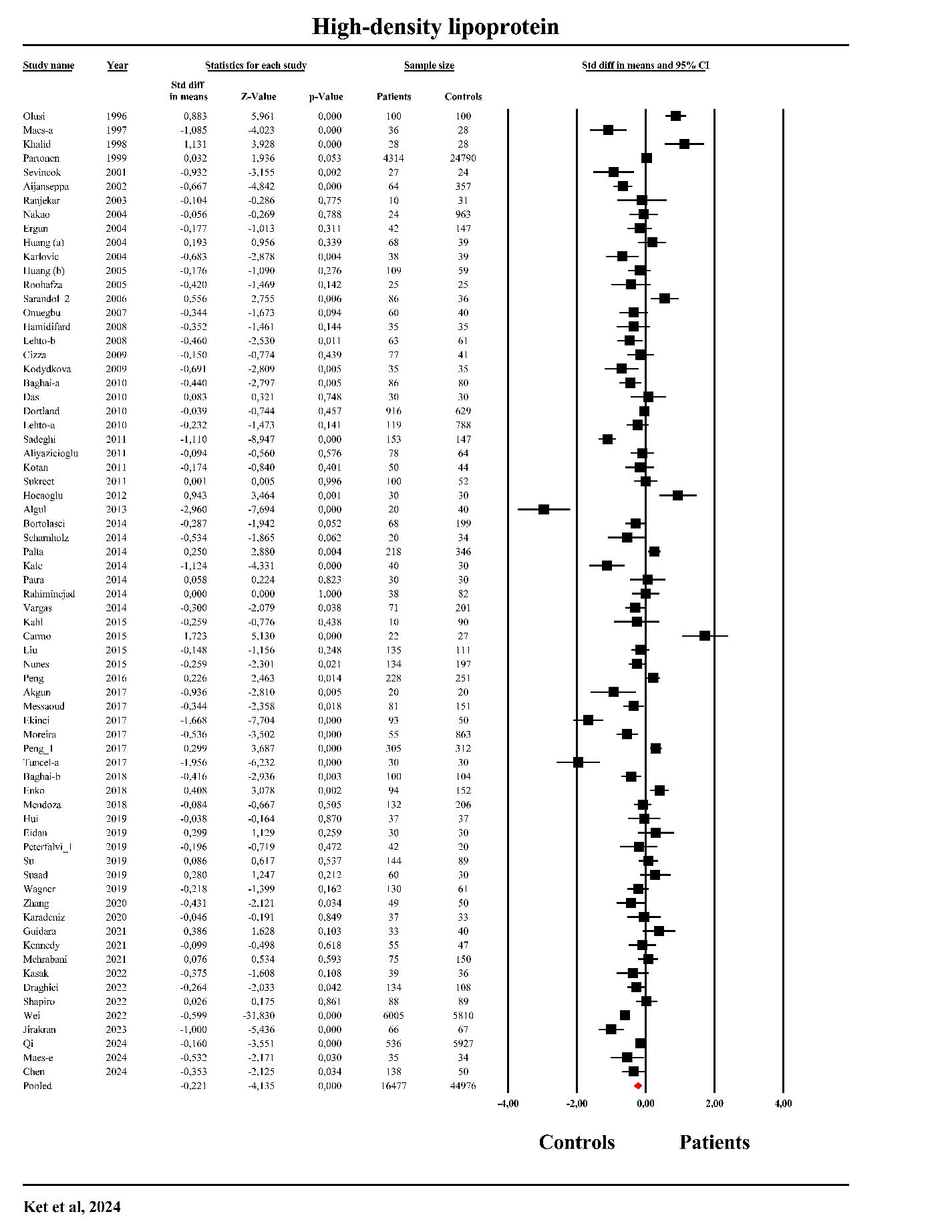

**ESF, Figure 8.** The forest plot of High-density lipoprotein (HDL) in patients with major depression as compared to healthy controls.

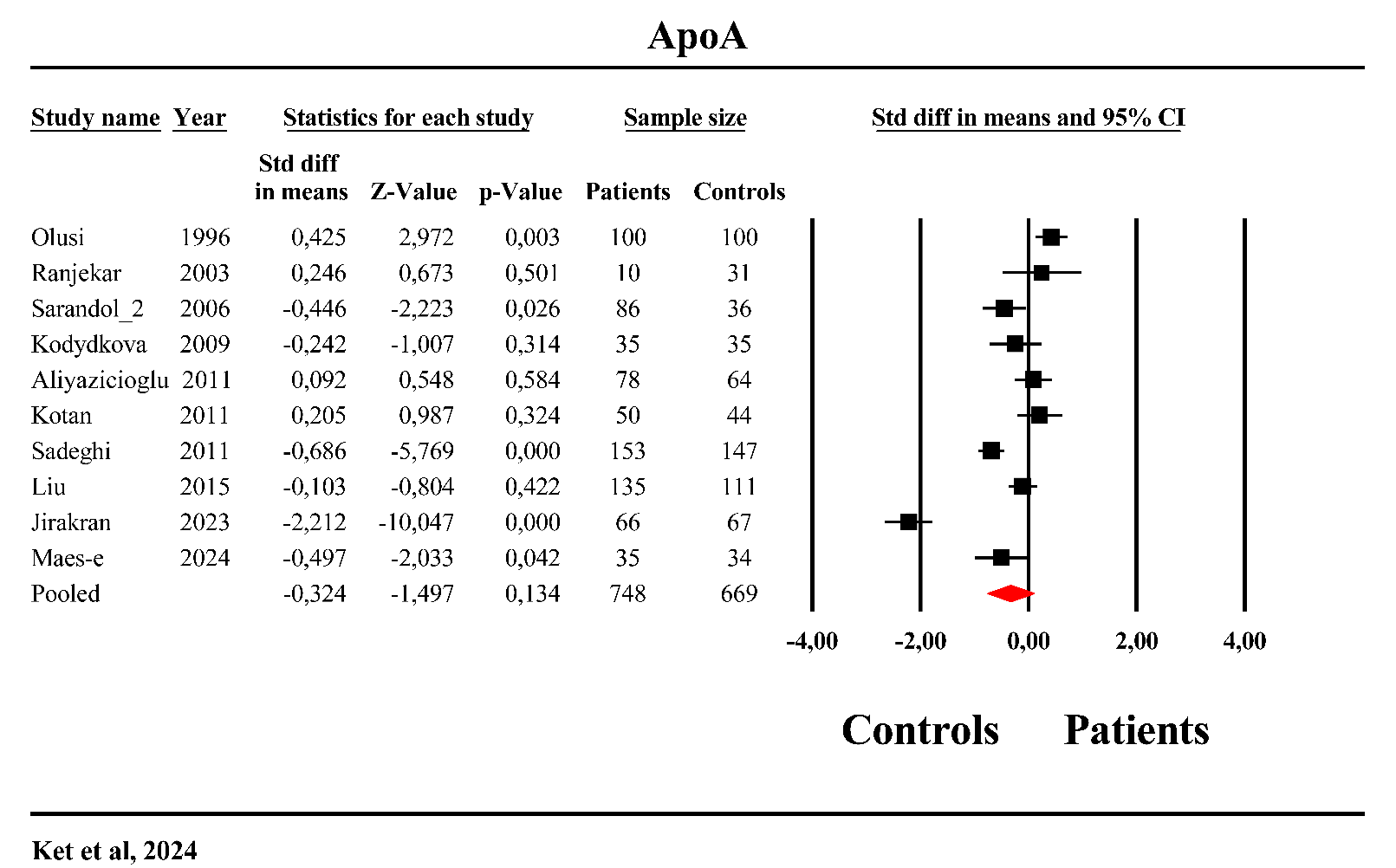

**ESF, Figure 9.** The forest plot of apolipoprotein (Apo)A in patients with major depression as compared to healthy controls.

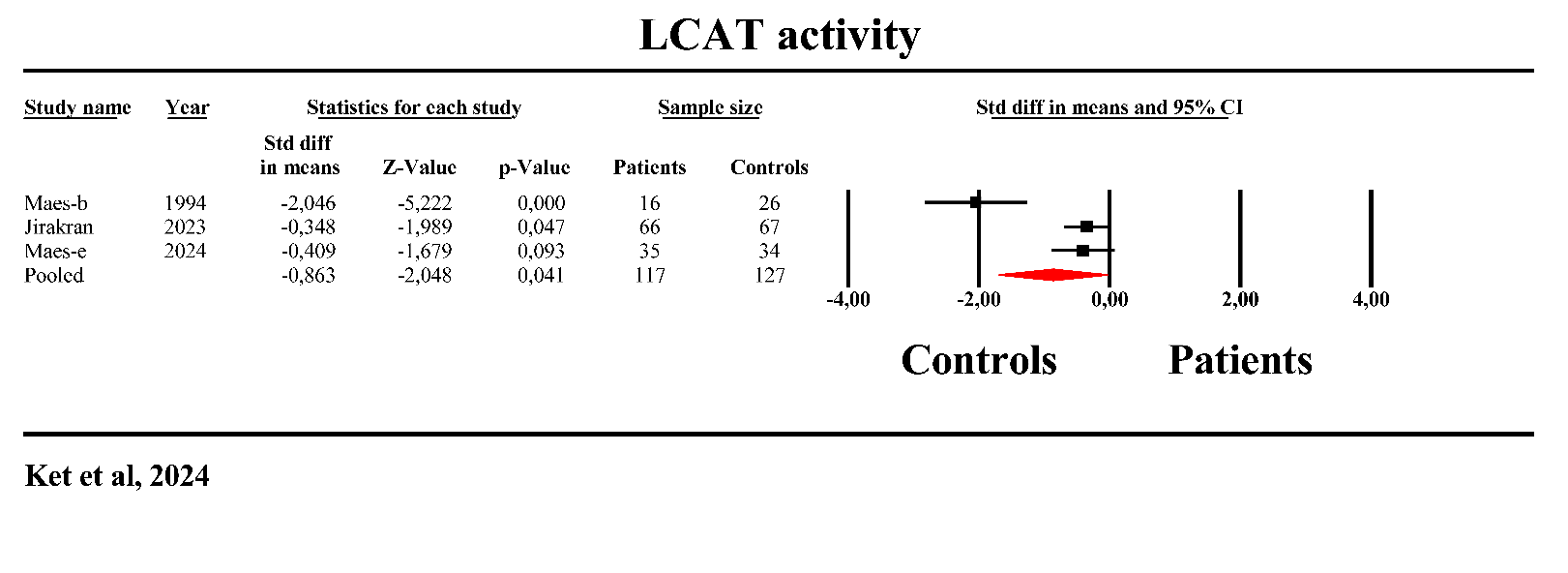

**ESF, Figure 10.** The forest plot of Lecithin cholesterol acyltransferase (LCAT) in patients with major depression as compared to healthy controls.

Chen, Q., T. Sun, Q. He, J. Yu, X. Zhang, L. Han and Y. Ren (2024). "Study of decreased serum levels of C1q/TNF-related protein 4 (CTRP4) in major depressive disorder." Journal of Psychiatric Research **172**: 274-280.

Khalfan, A. F., S. C. Campisi, R. F. Lo, B. W. McCrindle and D. J. Korczak (2023). "The association between adolescent depression and dyslipidemia." Journal of Affective Disorders **338**: 239-245.

Maes, M., B. Zhou, K. Jirakran, A. Vasupanrajit, P. Boonchaya-Anant, C. Tunvirachaisakul, X. Tang, J. Li and A. F. Almulla (2024). "Towards a major methodological shift in depression research by assessing continuous scores of recurrence of illness, lifetime and current suicidal behaviors and phenome features." J Affect Disord **350**: 728-740.

Qi, X., S. Wang, Q. Huang, X. Chen, L. Qiu, K. Ouyang and Y. Chen (2024). "The association between non-high-density lipoprotein cholesterol to high-density lipoprotein cholesterol ratio (NHHR) and risk of depression among US adults: A cross-sectional NHANES study." Journal of Affective Disorders **344**: 451-457.

Xu, K., S. Zhao, Y. Ren, Q. Zhong, J. Feng, D. Tu, W. Wu, J. Wang, J. Chen and P. Xie (2024). "Elevated SCN11A concentrations associated with lower serum lipid levels in patients with major depressive disorder." Transl Psychiatry **14**(1): 202.
